# Multi-model Diffusion MRI Signatures in Atypical Parkinsonian Disorders

**DOI:** 10.64898/2026.02.05.26345656

**Authors:** Yuqi Tian, Farwa Ali, Mary M. Machulda, Keith A. Josephs, Jennifer L. Whitwell

**Affiliations:** Department of Radiology, Mayo Clinic, Rochester, MN, USA; Department of Neurology, Mayo Clinic, Rochester, MN, USA; Department of Psychiatry and Psychology, Mayo Clinic, Rochester, MN, USA

**Keywords:** corticobasal syndrome, progressive supranuclear palsy-Richardson syndrome, Parkinson’s disease, Atypical parkinsonian syndromes, Diffusion MRI, NODDI, FBA, DTI, Free water DTI, Tissue weighted NODDI

## Abstract

Distinguishing atypical parkinsonian disorders (APS) from Parkinson’s disease (PD) remains challenging due to overlapping clinical features, yet accurate differentiation is critical for prognosis and treatment. Here, we employed multi-model diffusion MRI (dMRI) analysis to characterize microstructural alterations across corticobasal syndrome (CBS), progressive supranuclear palsy-Richardson syndrome (PSP-RS) and PD, with the aim of identifying which dMRI model provides optimum differentiation. We analyzed 25 CBS, 42 PSP-RS, and 21 PD participants compared to 35 age and sex-matched controls. Using a clinically feasible 3-shell high angular resolution diffusion imaging (HARDI) protocol, we applied 11 metrics from five complementary dMRI models—diffusion tensor imaging (DTI), free-water-eliminated model of DTI (FWE), neurite orientation dispersion and density imaging (NODDI), tissue-weighted NODDI, and Fixel Density (FD) in fixel-based analysis (FBA) —to comprehensively assess regional white and gray matter integrity. Group differentiation was assessed using Cohen’s d effect sizes and spearman correlations were assessed between dMRI metrics and clinical scales. Distinct microstructural signatures were observed across disorders and the sensitivity of the dMRI models differed. In group contrasts, DTI and NODDI-derived metrics consistently captured the strongest effects in midbrain and peduncular pathways for PSP-RS, whereas precentral and corticospinal alterations in CBS were most prominent using NODDI and FBA measures. Free-water-corrected metrics showed attenuated group differences. Across clinical-diffusion analyses, NODDI metrics exhibited the most robust associations with disease severity, while DTI and FWE measures detected more limited, regionally constrained effects. Together, these findings highlight complementary yet distinct sensitivities of tensor, free-water, multi-compartment, and fixel-based models to APS-related neurodegeneration.

## Introduction

Atypical parkinsonian syndromes (APS) comprise a group of neurodegenerative disorders that progress more rapidly and respond less favorably to levodopa than Parkinson’s disease (PD) (Testa, Monza et al. 2001). Among APS, progressive supranuclear palsy (PSP) and corticobasal syndrome (CBS) are the two most common and best-characterized syndromes (Litvan, Agid et al. 1996, Armstrong, Litvan et al. 2013, Hoglinger, Respondek et al. 2017). PSP-Richardson syndrome (PSP-RS), the classic PSP phenotype, is marked by early vertical supranuclear gaze palsy, axial rigidity, and postural instability with falls (Hauw, Daniel et al. 1994, Litvan, Agid et al. 1996), corresponding to degeneration of the midbrain and superior cerebellar peduncle (SCP) (Whitwell, Hoglinger et al. 2017). CBS presents with asymmetric cortical and subcortical symptoms—including limb apraxia, dystonia, and myoclonus—linked to degeneration of the frontal and parietal lobes (Boxer, Geschwind et al. 2006, Josephs, Whitwell et al. 2008). PD presents with resting tremor, rigidity,bradykinesia and postural instability, underpinned by α-synuclein pathology in the nigrostriatal system (Varadi 2020). In contrast, PSP-RS and CBS most commonly result from a 4-repeat tauopathy (Josephs, Hodges et al. 2011, Jabbari, Holland et al. 2020). Although PD, PSP-RS, and CBS share overlapping motor features, they target different neuroanatomical systems and exhibit distinct patterns of microstructural pathology and accurate differentiation is critical for prognosis and treatment.

Conventional diffusion tensor imaging (DTI) metrics such as fractional anisotropy (FA) and mean diffusivity (MD) provide a first-order characterization of tissue integrity (Basser and Pierpaoli 1996, Le Bihan, Mangin et al. 2001). Previous DTI studies have typically shown markedly reduced FA and increased MD in the SCP, midbrain, and frontal white matter in PSP-RS (Knake, Belke et al. 2010, Whitwell, Avula et al. 2011) (Canu, Agosta et al. 2011), and the callosal, thalamic, and frontoparietal tracts in CBS (Borroni, Garibotto et al. 2008, Boelmans, Kaufmann et al. 2009) (Chen, Liu et al. 2025). However, DTI is limited in specificity (Alexander, Hasan et al. 2001, Jones and Cercignani 2010, Jones, Knosche et al. 2013), as it cannot separate neuronal loss, demyelination, and free-water contamination. Several different diffusion MRI (dMRI) models have been developed to address these limitations. Free-water elimination (FWE) modeling improves sensitivity to intracellular diffusion changes by accounting for extracellular fluid, which may be particularly relevant for regions prone to cerebrospinal fluid (CSF) partial-volume effects such as the midbrain (Pasternak, Sochen et al. 2009, Pasternak, Westin et al. 2012). Neurite orientation dispersion and density imaging (NODDI) further partitions the diffusion signal into intra-cellular (ICVF) reflecting neurite density, isotropic (IsoVF) indexing free water, and orientation dispersion index (ODI) capturing the angular variability of neurite organization, offering more direct markers of axonal and dendritic microstructure (Zhang, Schneider et al. 2012). Complementarily, fixel-based analysis (FBA) quantifies fiber-specific density (FD, featuring microstructures) within crossing-fiber regions, allowing detection of tract-specific degeneration that may underlie focal symptoms (Raffelt, Smith et al. 2015, Raffelt, Tournier et al. 2017).

By applying these advanced dMRI models within a unified statistical framework, we aimed to determine which diffusion metrics best differentiate PSP-RS and CBS from controls and PD, and to identify disease-specific neuroanatomical signatures. We also aimed to evaluate correlations between diffusion metrics and clinical measures of disease severity to help evaluate the value of these metrics as disease biomarkers. To our knowledge, this is the first study to systematically evaluate the ability of different dMRI models to differentiate between these parkinsonian syndromes, integrating pairwise disease contrasts and clinical-diffusion associations. We hypothesized that advanced dMRI models would demonstrate superior effect sizes and clinical relevance compared with conventional DTI.

## Methods

### Participant Recruitment

Participants were prospectively recruited by the Neurodegenerative Research Group (NRG) from the Department of Neurology, Mayo Clinic, Rochester, MN, between 2018 and 2025 as part of ongoing studies of APS. Twenty-five participants who met clinical criteria for possible or probable CBS (Armstrong, Litvan et al. 2013) and 42 participants who met clinical criteria for probable PSP-RS (Litvan, Agid et al. 1996, Hoglinger, Respondek et al. 2017) were recruited and compared to 21 participants with PD and 35 cognitively unimpaired controls.

Participants with CBS who had biomarker evidence of Alzheimer’s disease, i.e. both beta-amyloid and temporal lobe tau deposition observed on Positron Emission Tomography (PET), were excluded from the study. Participants with PSP-RS who had clinical features of CBS were excluded. The controls were cognitively and motorically normal participants who did not have any complaints of cognitive, motor, or behavioral abnormalities and had a score of ≥23 on the Montreal Cognitive Assessment battery (MoCA) (Nasreddine, Phillips et al. 2005) and a score of 0 on the Hoehn and Yahr scale (Hoehn and Yahr 1967). All participants underwent the same standardized research 3T MRI protocol performed on a Siemens scanner.

The participants with CBS, PSP-RS, and PD were evaluated by movement disorder specialists (FA or KAJ). Comprehensive assessments included the MoCA (Nasreddine, Phillips et al. 2005), the Movement Disorder Society sponsored revision of the Unified Parkinson’s Disease Rating Scale Part III (MDS-UPDRS III) (Goetz, Tilley et al. 2008), PSP Rating Scale (Golbe and Ohman-Strickland 2007), Frontal Assessment Battery (FAB) (Dubois, Slachevsky et al. 2000), Western Aphasia Battery Praxis subtest (WAB-Praxis) (Shewan and Kertesz 1980), PSP Saccadic Impairment Scale (PSIS) (Whitwell, Master et al. 2011), and the Test of Upper Limb Apraxia (TULIA) (Vanbellingen, Kersten et al. 2010), including left- and right-hand subscores when applicable. Disease-specific onset variables such as age at onset and duration from symptom onset to examination were also collected.

All participants provided written informed consent under protocols approved by the Mayo Clinic Institutional Review Board.

### Image processing

All participants underwent a standardized multi-shell high angular resolution diffusion imaging (HARDI) protocol on a 3T Siemens Prisma scanner (Siemens Healthineers, Erlangen, Germany) using a 64-channel head/neck coil. Diffusion-weighted images (DWI) were acquired using a multiband-accelerated (MB=3) single-shot echo-planar imaging (EPI) sequence (ep2d_diff), with b = 0, 500, 1000, and 2000 s/mm², monopolar diffusion encoding, TE = 71 ms, TR = 3.4 s, slice thickness = 2 mm, and an in-plane matrix of 116 × 116 (voxel size 2 × 2 × 2 mm³).

Diffusion data was preprocessed using a standard FSL/ MRtrix3 pipeline. Steps included: denoising, Gibbs ringing removal, motion and eddy-current correction with FSL eddy (including outlier replacement), and bias-field correction (ANTS-N4). A brain mask was generated with MRtrix3, and all gradient tables were verified after eddy correction. Preprocessed data were visually inspected for artifacts, distortion correction, and alignment across shells before downstream tensor fitting and microstructural modeling.

### Diffusion Models

Following preprocessing and atlas registration, 4D DWI volumes were analyzed using multiple diffusion models to capture complementary microstructural features.

**DTI:** A weighted least-squares tensor fit (DIPY(Garyfallidis, Brett et al. 2014)) was applied to derive FA and MD from the tensor eigenvalues. **FWE:** Free-water elimination (DIPY) was used to decompose the signal into tissue and extracellular compartments, yielding free-water-corrected FA and MD. **NODDI:** The three-compartment NODDI model was fitted using AMICO (Daducci, Canales-Rodriguez et al. 2015) to obtain ICVF, ODI, and IsoVF. **FBA:** Using MRtrix3, fiber orientation distributions were estimated with constrained spherical deconvolution (lmax = 8). Fixel-wise FD was extracted as the primary microstructural metric. Quality control was performed at each reconstruction stage to ensure model stability and physiological plausibility.

For NODDI measures, we additionally computed tissue-weighted means to reduce bias from CSF partial-volume contamination (Parker, Veale et al. 2021). Instead of a simple arithmetic mean: 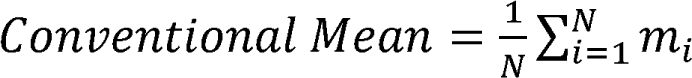 we applied a tissue-fraction-weighted mean: Tissue-Weighted 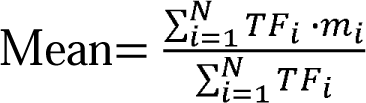 where *m_i_* is the metric value (e.g., ICVF) and TF_i_ is the tissue fraction estimated by the NODDI model in voxel *i*. This approach reduces the influence of voxels with substantial CSF contribution.

DTI metrics use single-tensor fitting and do not incorporate tissue-fraction estimates. FBA metrics are derived from fixels and inherently model partial-volume effects across tissue types; therefore, additional tissue-weighted averaging is not required for FBA.

### Region of Interest (ROI) extraction

Diffusion metrics from all reconstruction models were extracted using the JHU EVE white matter atlas (Oishi, Faria et al. 2009), which was registered to each participant’s FA image and restricted to voxels within the DWI brain mask. ROI-wise means were computed for DTI, FWE-DTI, NODDI, tissue-weighted NODDI, and FBA metrics across all voxels within each region. Metrics were extracted separately for left and right hemispheric ROIs when applicable, and bilateral values were defined as the arithmetic mean of the two sides. ROIs with insufficient coverage (<50% of voxels containing valid diffusion data) were excluded. All ROI maps were visually inspected to confirm accurate registration and to identify any regions affected by artifacts.

### Statistical Analysis

All statistical analyses were performed in Python using the pandas, scipy, statsmodels, and matplotlib packages. Regional dMRI metrics were extracted from 118 predefined ROIs in the JHU Type III white matter atlas.

Group effects across four diagnostic categories (CBS, PD, PSP-RS and controls) were evaluated using Analysis of Covariance (ANCOVA), with diagnosis as the main factor and age, sex, and disease duration included as covariates when applicable. For each ROI × metric, we obtained omnibus p-values and partial eta-squared (η²) as a measure of effect size. Pairwise post-hoc contrasts between diagnostic groups were derived from the ANCOVA model, yielding adjusted mean differences and p-values for each pairwise comparison. Effect sizes for pairwise contrasts were expressed as Cohen’s d, computed from group-specific adjusted means and the pooled residual variance from the ANCOVA model.

To control for multiple comparisons across ROIs, we applied two procedures within each diffusion metric: (i) Bonferroni correction, yielding family-wise error–corrected p values (pval_fwe), which served as the primary criterion for statistical significance; and (ii) the Benjamini–Hochberg false discovery rate (FDR) procedure, yielding FDR-corrected q values (q), which are reported for reference. Both omnibus and pairwise ANCOVA results are reported with cohen’s d, raw p values, q, and pval_fwe.

Partial Spearman correlations were computed between regional diffusion metrics and clinical scores, controlling for age, sex, and disease duration when applicable. For each metric-score pair, we report Spearman’s ρ, uncorrected p value, sample size n, FDR-corrected q value, and Bonferroni-corrected pval_fwe, with correction applied separately for each clinical score across all ROI × metric combinations.

All statistical tests were two-sided, and q < 0.05 was considered statistically significant.

### Data Availability

The data that supports the findings of this study are available from the corresponding author, upon reasonable request.

## Results

### Clinical characteristics

The demographic and clinical characteristics of the cohort are summarized in **Table 1**. Years of education and handedness did not differ across all groups or within disease groups, whereas the proportion of female participants and age at encounter differed across groups. The time from symptom onset to visit differed across disease groups, and was longest in PD. As expected, multiple clinical scales demonstrated disease-related impairments. PSP-RS showed the lowest MoCA and FAB scores and the highest MDS-UPDRS III, PSP Rating Scale, PSP Rating scale gait/midline, and PSIS scores. CBS exhibited the most pronounced deficits on praxis measures, with the lowest WAB-Praxis and TULIA scores. Left- and right-hand TULIA subscores similarly differed across participant groups. Post hoc pairwise comparisons with multiple-comparison correction were performed (see continuous results in **Table S1** and categorical results in **Table S2**), with sample sizes based on non-missing observations reported for each group. Together, these results confirm the expected cognitive, motor, and praxis impairments characteristic of CBS, PD, and PSP-RS.

**Table 1.** Demographic and clinical characteristics of study participants. Data is median (IQR) for continuous variables and n (%) for categorical variables. Group differences were assessed using the Kruskal-Wallis test (continuous) and chi-square or Fisher’s exact test (categorical). Clinical scales: MoCA (Montreal Cognitive Assessment); FAB (Frontal Assessment Battery); MDS-UPDRS III (Movement Disorder Society-sponsored revision of the Unified Parkinson’s Disease Rating Scale, Part III); PSP Rating Scale (Progressive Supranuclear Palsy Rating Scale); PSP Rating Scale gait/midline score (Progressive Supranuclear Palsy Rating Scale gait/midline score); WAB-Praxis (Western Aphasia Battery - Praxis Subtest); TULIA total (Test of Upper Limb Apraxia); TULIA-Left and TULIA-Right (left/right subscores); PSIS (PSP Saccadic Impairment Scale).Statistical significance was set at p<0.05.

| Variable | Control (n=35) | CBS (n=25) | PD (n=21) | PSP-RS (n=42) | P across all groups | P across CBS, PSP-RS and PD |
| --- | --- | --- | --- | --- | --- | --- |
| Age at visit, years | 66 (63, 70) | 68 (64, 72) | 69 (66, 78) | 72 (68, 75) | 0.008 | 0.08 |
| Age at onset, years | N/A | 65 (60, 68) | 64 (59, 66) | 68 (63, 72) | N/A | 0.03 |
| Time from onset to visit, years | N/A | 3 (2, 4) | 5 (4, 8) | 3 (3, 5) | N/A | 0.0003 |
| Education, years | 16 (15, 16) | 14 (12, 16) | 16 (12, 16) | 14 (12, 16) | 0.17 | 0.86 |
| Female sex, N (%) | 18 (51%) | 15 (60%) | 5 (24%) | 13 (31%) | 0.02 | 0.08 |
| Handedness - left or ambidextrous | 3 (9%) | 1 (4%) | 1 (5%) | 5 (12%) | 0.64 | 0.61 |
| Moca (/ 30) | 27 (26, 28) | 23 (20, 24) | 25 (23, 27) | 22 (19, 25) | <0.001 | 0.02 |
| MDS-UPDRS III (/ 132) | 1 (0, 2) | 33 (23, 54) | 23 (17, 44) | 37 (30, 52) | <0.001 | 0.05 |
| PSP Rating Scale (/ 100) | N/A | 26 (21, 40) | 12 (7, 18) | 36 (28, 40) |  | <0.001 |
| PSP Rating Scale gait/midline score (/ 20) | N/A | 10 (5, 14) | 4 (1, 7) | 11 (7, 15) |  | <0.001 |
| FAB (/ 18) | N/A | 14 (12, 17) | 16 (14, 17) | 13 (11, 14) |  | 0.0008 |
| WAB_Praxis (/ 60) | 60 (58, 60) | 52 (49, 54) | 60 (60, 60) | 56 (56, 56) | <0.001 | <0.001 |
| PSIS (/ 5) | 0 (0, 0) | 1 (0, 2) | 0 (0, 1) | 3 (2, 4) | <0.001 | <0.001 |
| TULIA (/ 240) | 12 (11, 12) | 5 (2, 9) | 12 (12, 12) | 11 (10, 12) | <0.001 | <0.001 |
| TULIA_left (/ 12) | 12 (11, 12) | 9 (7, 10) | 12 (12, 12) | 11 (10, 12) | <0.001 | <0.001 |
| TULIA_right (/ 12) | 12 (11, 12) | 10 (5, 11) | 12 (12, 12) | 12 (10, 12) | 0.001 | 0.0008 |

### Group comparisons

Whole-brain ROI-wise ANCOVA and post-hoc pairwise comparisons identified the ROI-metric combinations that best differentiated CBS from controls and PD (**Figure 1**), PSP-RS from controls and PD (**Figure 2**), and CBS from PSP-RS (**Figure 3**). Each figure displays the top 50 ROI-metric pairs ranked by effect size, with family-wise error-corrected p-values shown in the cells. The PD vs Control comparison is presented in Supplemental **Figure S1**. The tables with detailed data are Supplemental **Table S3 - S8**.

**Figure 1.**
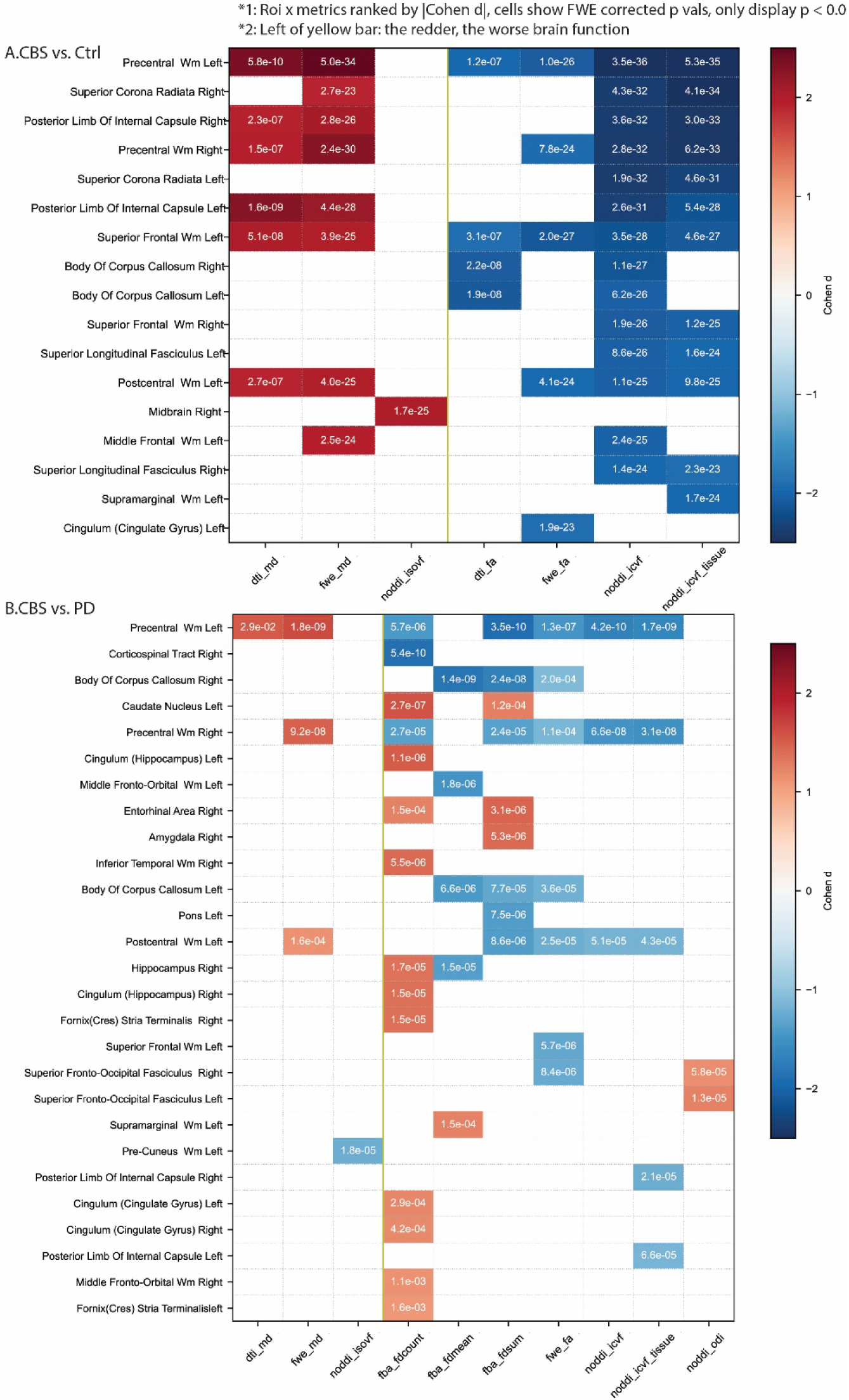
Top 50 ROI-metric pairs ranked by |Cohen’s d| for each group comparison. (A) CBS vs Controls. Cell colors represent the magnitude of the effect size (|Cohen’s d|), and each cell displays pval_fwe. Metrics are arranged by polarity, with the left of the yellow midline indicating metrics for which higher values reflect worse brain integrity relative to controls.(B) CBS vs PD. Visualization is identical to panel A, showing the top 50 ROI-metric pairs ranked by |Cohen’s d| for the comparison. Distinguishing features cluster in precentral and corticospinal pathways, prominently captured by FWE-MD, NODDI-ICVF, ICVF-tissue, and FBA-FD measures.

**Figure 2.**
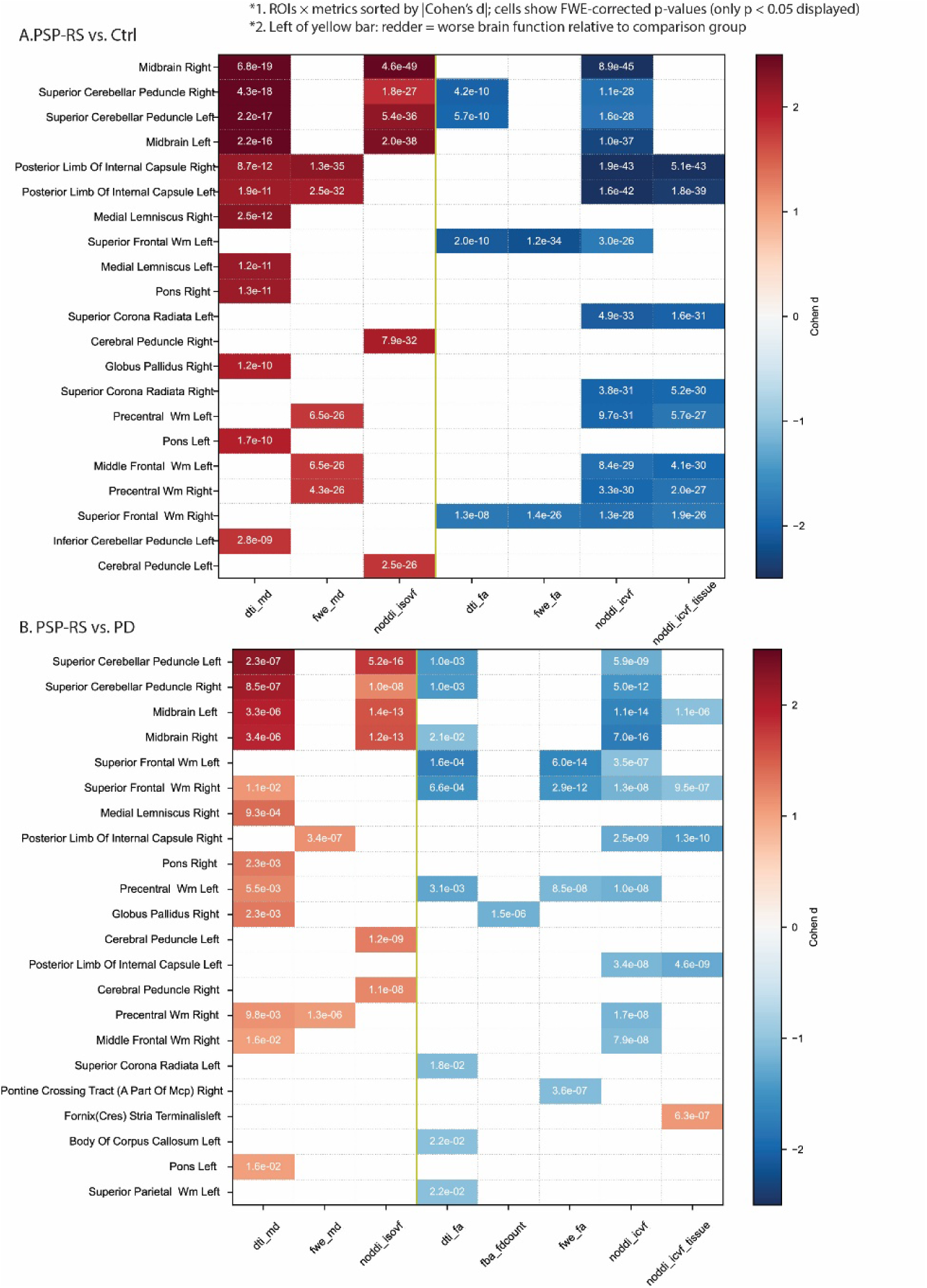
Syndrome-specific microstructural differences of PSP-RS relative to controls and PD. Top 50 ROI-metric pairs ranked by |Cohen’s d| for PSP-RS vs controls (A) and PSP-RS vs PD (B). **A**. PSP-RS vs controls: Prominent abnormalities localize to brainstem motor pathways, including the midbrain, SCP, and PLIC, with NODDI IsoVF and ICVF and DTI MD showing the largest effect sizes. **B**. PSP-RS vs PD: Key differences localize to the bilateral midbrain and SCP, with strongest separation in DTI-MD, NODDI IsoVF, and NODDI-ICVF. NODDI metrics show consistent sensitivity across both contrasts.

**Figure 3.**
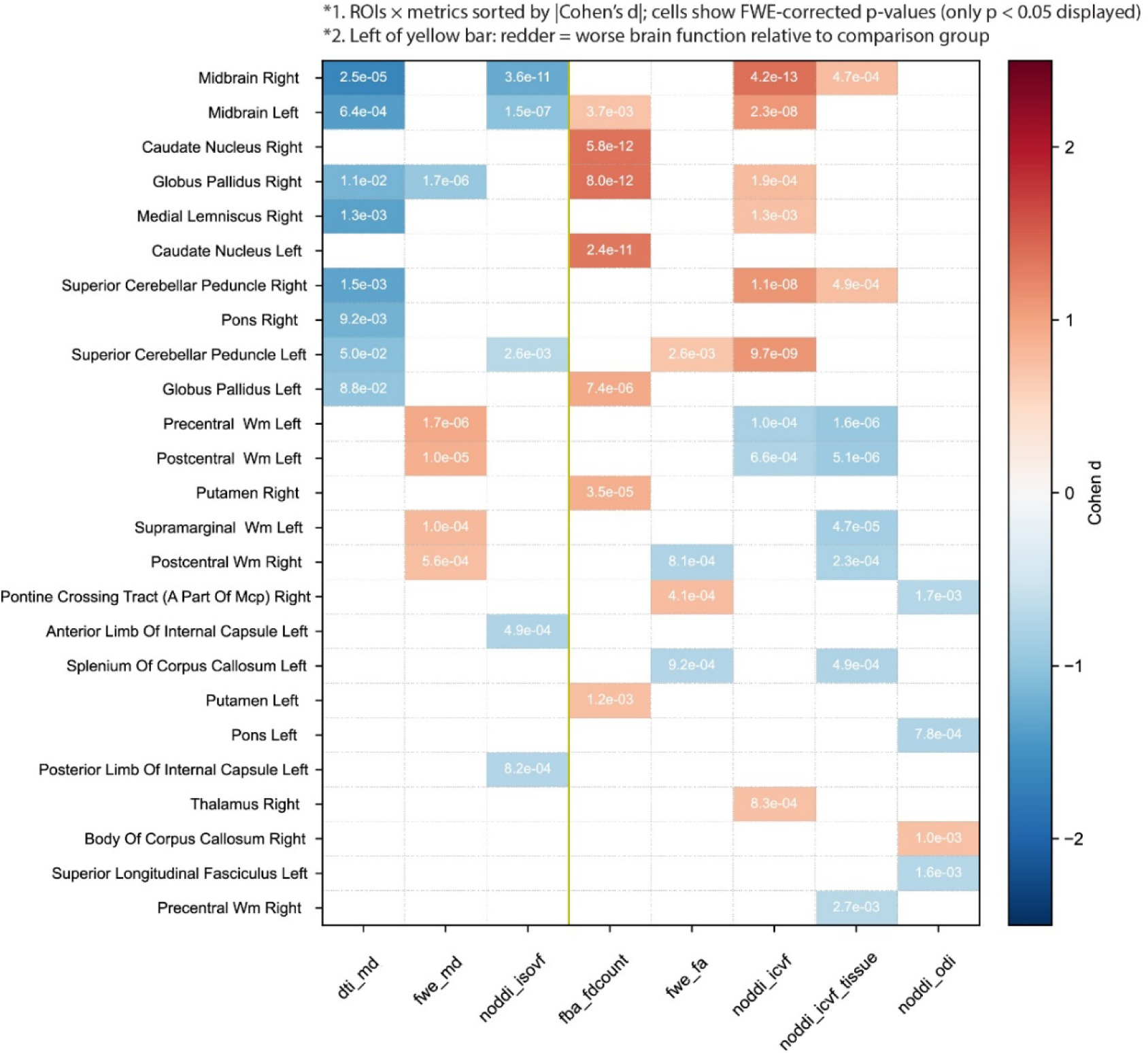
Syndrome-specific microstructural differences of CBS relative to PSP-RS. Top 50 ROI-metric pairs ranked by |Cohen’s d| for CBS vs PSP-RS. Largest effect sizes were observed in the bilateral midbrain, caudate, and globus pallidus, primarily for DTI, NODDI (ICVF, IsoVF), and FBA metrics.

#### CBS versus controls and PD

The most prominent CBS-related abnormalities compared to controls (**Figure 1A**) localized to the bilateral precentral cortex, where FWE MD, DTI MD, NODDI ICVF, and NODDI ICVF tissue showed the largest effect sizes (|cohen’s d| > 1.9) accompanied by significant pval_fwe labelled in the cells. Additional abnormalities were observed in the superior corona radiata (SCR), posterior limb of the internal capsule (PLIC), and superior frontal and postcentral white matter, with NODDI metrics demonstrating greater sensitivity than FWE or DTI in both magnitude and spatial extent.

When contrasted with PD (**Figure 1B**), a broader distribution of metrics and ROIs were identified in the top 50. CBS remained primarily distinguished by worse integrity of the precentral cortex, with the highest cohen’s d values observed for DTI MD, FWE MD, FBA FD count and sum, NODDI ICVF and ICVF tissue. The corticospinal tract and body of the corpus callosum showed high cohen’s d values for the FBA measures, as well as FWE FA for the body of the corpus callosum. The FBA FD sum, FWE FA and NODDI measures also identified abnormalities in the postcentral white matter, with the PLIC identified by NODDI. The FBA measures also identified several ROIs with worse integrity in PD, including the cingulum and temporal lobe ROIs.

#### PSP-RS versus controls and PD

The most prominent PSP-RS abnormalities compared to controls (**Figure 2A**) were observed within brainstem motor pathways, including the midbrain, SCP, and PLIC. NODDI IsoVF and ICVF, and DTI MD, produced the highest effect sizes in these regions, outperforming FWE measures. NODDI-ICVF and ICVF-tissue also revealed abnormalities extending into the SCR, middle and superior frontal white matter (SFWM) and precentral white matter. These frontal white matter regions were also identified by FWE MD and FA.

When contrasted with PD (**Figure 2B**), the largest effect sizes occurred in the bilateral SCP and midbrain with DTI-MD, NODDI IsoVF, and NODDI ICVF all showing strong discriminative performance. Abnormalities in PSP-RS were also identified in frontal and precentral white matter ROIs by DTI MD and FA, FWE MD and FA, NODDI ICVF and NODDI ICVF tissue. Across most of the top-ranked ROIs, PSP-RS showed greater microstructural impairment than PD. One exception was the left fornix/stria terminalis, where PSP-RS demonstrated higher NODDI ICVF tissue than PD—a pattern consistent with more severe fornix degeneration in PD in this cohort.

#### CBS versus PSP-RS

**Figure 3** shows that large effect sizes in the bilateral midbrain, right globus pallidus, right medial lemniscus, and SCP were captured by both DTI MD and NODDI ICVF, whereas their free-water-corrected counterparts exhibited attenuated effects. Across the top-ranked ROIs, polarity consistently indicated lower free-water diffusion (DTI MD), higher neurite density (NODDI ICVF), and higher fixel counts (FBA FD count) in CBS compared with PSP-RS, suggesting worse microstructural integrity in PSP-RS within these regions. Notable exceptions to this pattern included the precentral cortex, supramarginal white matter, postcentral white matter, and the splenium of the corpus callosum, which showed worse integrity in CBS and were captured by FWE MD, FWE FA, NODDI ICVF, NODDI ICVF tissue, and NODDI ODI. Within the pons, DTI MD identified lower free-water diffusion in the right pons, whereas NODDI ODI detected reduced dispersion in the left pons among the top 50 ROI × metric effect sizes, highlighting complementary model sensitivity. High effect sizes in the bilateral caudate nucleus were detected exclusively by FBA, highlighting worse integrity in PSP-RS.

#### Analysis of key ROIs

**Figure 4 A-F** shows the direct comparison of effect sizes for the metrics in key regions including the precentral cortex, SCP, and midbrain bilaterally. **Supplemental Figure S2-7** displays the absolute values with letter codes showing FDR-corrected q values across 3 syndromes and controls for all metrics at left and right key ROIs. For the precentral cortex in CBS, the DTI, FWE, and NODDI ICVF metrics all showed high effect sizes in comparison to controls and PD, with the highest values observed with NODDI ICVF, although FBA FD-sum also showed a high effect size in comparison to PD (**Figure 4A and B**). In comparison to PSP-RS, the effect sizes for the precentral cortex were relatively low, and only FWE (MD and FA) and NODDI ICVF differed between PSP-RS and CBS (**Figures 4C**). For the midbrain and SCP in PSP-RS, the highest effect size was found with DTI MD in comparison to controls, PD and CBS, with the next highest effect sizes observed for NODDI IsoVF and ICVF. Notably, the FBA metrics were not sensitive to abnormalities in the SCP in PSP-RS (**Figure 4D and E**).

**Figure 4.**
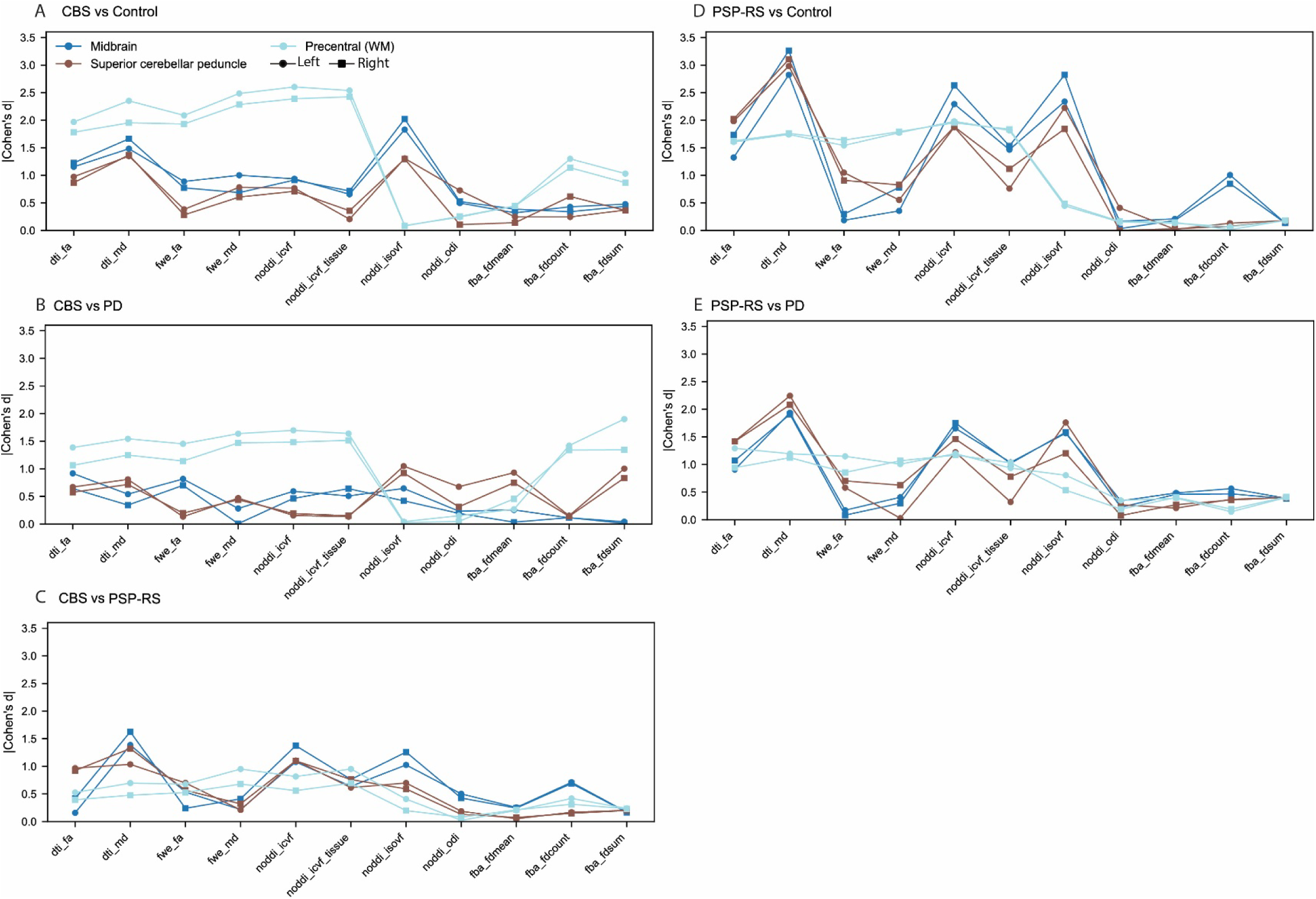
Direct comparison of effect sizes for diffusion metrics across key regions: precentral cortex (light blue), superior cerebellar peduncle (SCP; brown), and midbrain (dark blue), bilaterally. Panels illustrate pairwise group contrasts: (A) CBS vs Controls, (B) CBS vs PD, (C) CBS vs PSP-RS, (D) PSP-RS vs Controls, (E) PSP-RS vs PD, and (F) PD vs Controls. Round markers indicate the left hemisphere; square markers indicate the right hemisphere.

### Clinical correlations

Spearman correlation analyses examined associations between diffusion metrics and clinical measures. **Figure 5** presents partial Spearman correlations adjusted for age, sex, and disease duration. The x-axis shows the rank order of Spearman’s rho values, and the y-axis displays -log10(pval_fwe). Only ROI-metric pairs with |rho| > 0.4 and pval_fwe < 0.05 are included. Comprehensive results for clinical measures in **Table 1** (MoCA, MDS-UPDRS III, PSP Rating Scale, PSP Rating Scale gait/midline score, FAB, WAB_praxis, PSIS, TULIA) across the whole cohort and within disease-specific subgroups are shown in Supplementary **Figure S8 - S22**.

**Figure 5.**
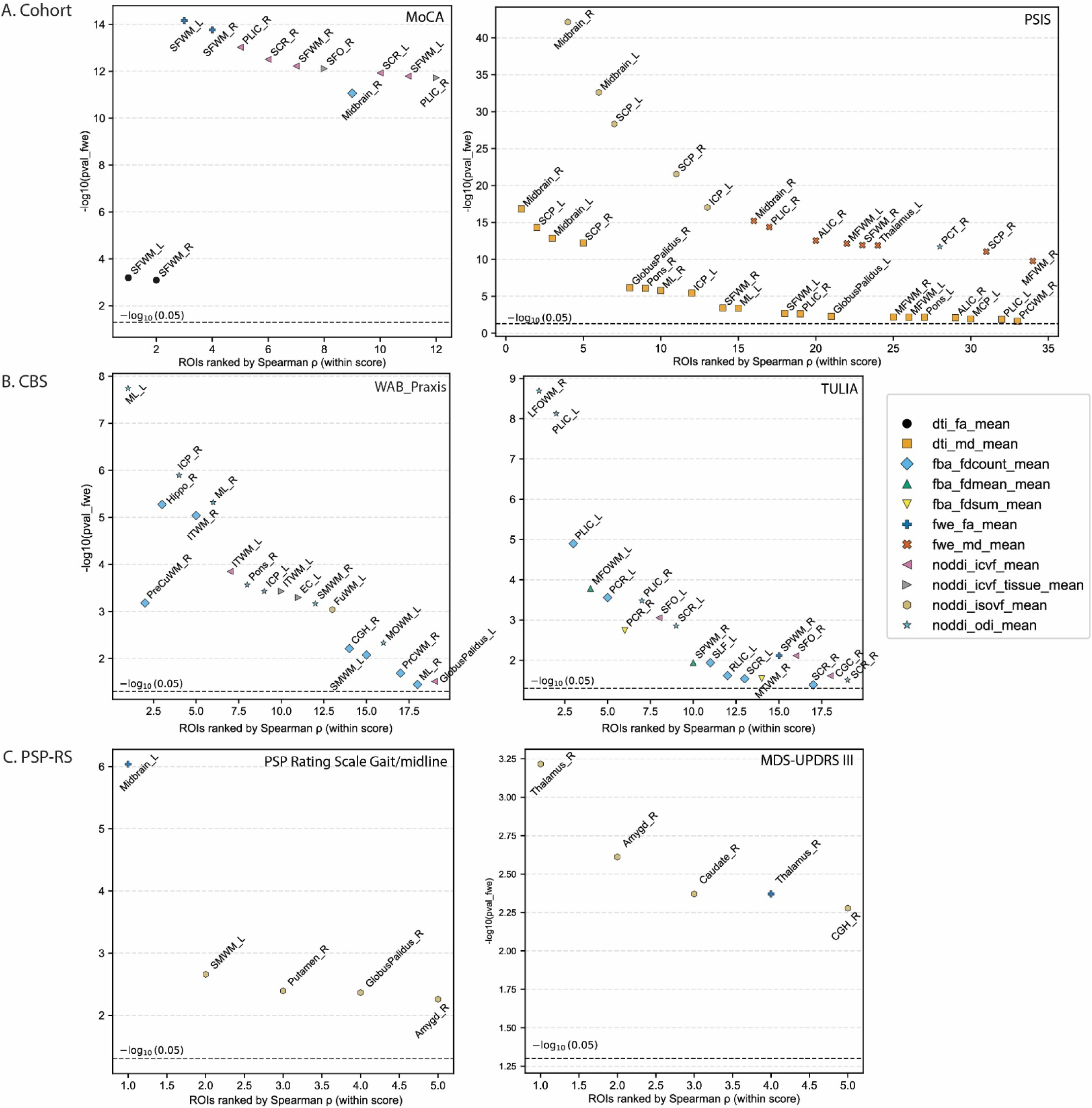
Diffusion-clinical correlation signatures across the cohort and within CBS and PSP-RS. Partial Spearman correlations (adjusted for age, sex, and disease duration) highlight distinct anatomical substrates of clinical impairment. A. Whole cohort: MoCA correlates with frontal white matter integrity, while PSIS correlates with brainstem pathways (midbrain, SCP). B. CBS: Praxis deficits (WAB-Praxis, TULIA) map to axonal dispersion and fiber-density loss in precentral, cerebellar, and temporoparietal tracts, primarily detected by NODDI-ODI and FBA-FD. C. PSP-RS: PSP Rating Scale Gait/midline and motor impairment (MDS-UPDRS III) correlate strongly with NODDI IsoVF in the midbrain, basal ganglia, and thalamus. Only ROI-metric pairs with |ρ| > 0.4 and pval_fwe < 0.05 are shown.

Given that cognitive impairment can be a feature of both CBS and PSP-RS, correlations with MoCA were assessed in the whole cohort (**Figure 5A**). Strongest correlations were observed in bilateral SFWM, with DTI FA showing the largest Spearman rho values but FWE-FA showing the smallest p values (-log10(pval_fwe) ≈ 14). Additional associations were observed in the right PLIC, right SCR and right SFWM for NODDI ICVF. Correlations with PSIS revealed a distinct brainstem signature, with significant associations in the bilateral midbrain and SCP. These were strongest for NODDI IsoVF (-log10(pval_fwe) > 20), followed by DTI MD and FWE MD. Within the PSP-RS subgroup, PSIS scores showed a restricted range with generally high values, and only a single ROI exhibited a correlation with |ρ| > 0.4, limiting interpretability at the subgroup level (**Figure S19**). Accordingly, whole-cohort analyses provided more informative and stable estimates of PSIS-diffusion relationships.

Within CBS (**Figure 5B**), correlations with WAB-Praxis were found in the medial lemniscus and right inferior cerebellar peduncle with NODDI ODI, as well as in the right precuneus white matter, right hippocampus, and right inferior temporal white matter using FBA FD. With TULIA, correlations were observed in the right lateral fronto-orbital white matter and bilateral PLIC using NODDI ODI, and in the left PLIC and left posterior corona radiata using FBA-FD.

Within PSP-RS (**Figure 5C**), the PSP Rating Scale-GM and MDS-UPDRS III correlations highlighted a brainstem-basal ganglia network: For PSP Rating Scale-GM, strong correlations emerged in the left midbrain using FWE-FA, and in left supramarginal white matter, right putamen, right globus pallidus, and right amygdala using NODDI IsoVF.. For MDS- UPDRS III, high correlations emerged in the right thalamus, right amygdala, right caudate, and right cingulum-hippocampal region, again predominantly captured by NODDI IsoVF.

## Discussion

Our findings reveal clear and biologically grounded differences in diffusion-derived microstructure across CBS, PSP-RS, and PD and support dMRI measures, particularly NODDI metrics, as potential diagnostic biomarkers in APS.

In CBS, optimum differentiation from controls was observed with the precentral white matter, SCR, PLIC, as well as the superior frontal and postcentral regions, using NODDI ICVF, ICVF tissue, FWE MD, and DTI MD. The FA measures from DTI and FWE and FBA measures also picked up differences in the precentral white matter but with lower effect sizes. However, NODDI IsoVF and NODDI ODI did not detect any abnormalities in the precentral white matter in CBS versus controls. These findings overlap with one study (Boelmans, Kaufmann et al. 2009), that reported FA reductions in the CST and Corpus Collosum trunk which are tracts that inherently pass through the precentral white matter, SCR, and PLIC, and partially align with another (Borroni, Garibotto et al. 2008), whose Corticobasal Degeneration Syndrome-related FA reductions in sensorimotor associative fibers and intraparietal-frontal pathways extend into regions adjacent to the precentral and postcentral cortices, but do not include the SCR or PLIC. The findings also indicate that ICVF is more sensitive than the other NODDI metrics, as it directly reflects neurite/axonal density, while ODI and FA are more influenced by fiber orientation complexity and partial volume effects. This same limitation likely explains why MD is more sensitive than FA. IsoVF, which reflects free extracellular water, may be less sensitive in these regions where early axonal loss predominates over fluid expansion.

¹□F□Fluorodeoxyglucose (FDG) PET work in CBS (Ghirelli, Goodrich et al. 2025) has demonstrated hypometabolism in parietal and perirolandic cortices with relative sparing of medial temporal and basal ganglia regions, a pattern that closely matches the diffusion-derived perirolandic and temporoparietal abnormalities observed here.

Compared with PD, CBS remained characterized by reduced integrity in the precentral white matter, again detected by the MD measures and NODDI ISOVF, although several other tracts were also identified, including the body of the corpus callosum and corticospinal tract, which were most strongly detected by the FBA measures. In addition, the FBA measures identified several regions which showed worse integrity in PD compared to CBS, including regions in the temporal lobe, cingulum and fornix. These findings may indicate the presence of underlying Alzheimer’ disease (AD) pathology in the PD cohort, since AD is associated with reduced integrity in these tracts. Concomitant AD in PD is common (Tropea, Albuja et al. 2023), with approximately 10% of PD without dementia and ∼35% of PD with dementia showing evidence of AD co pathology. Our tract based findings partially overlap with the significant hypometabolism observed in temporal lobe regions of concomitant AD in PD (Castro-Labrador, Silva-Rodríguez et al. 2024). Unfortunately, Aβ PET scanning was not performed in our PD cohort to allow us to test this hypothesis. In addition, the broader pattern identified by FBA in the CBS-PD comparison likely reflects its sensitivity to fiber-specific degeneration that is not captured by voxel-averaged diffusion metrics, such as FA and NODDI ODI.

As expected, the PSP-RS cohort showed different patterns of abnormalities, with reduced integrity in the midbrain, SCP and PLIC, consistent with previous DTI studies in PSP-RS (Knake, Belke et al. 2010, Canu, Agosta et al. 2011, Whitwell, Avula et al. 2011). Similar to the CBS comparisons, the DTI and FWE MD measures and NODDI ICVF were especially sensitive to these changes, although NODDI IsoVF also produced high effect sizes. Abnormalities were also observed in frontal white matter, predominantly with NODDI ICVF. Hence, across comparisons, NODDI metrics generally outperformed DTI and FWE, with larger effect sizes and clearer syndrome-specific patterns. These results support the value of multi-compartment modeling for revealing subtle but meaningful disease processes in APS. Consistent with the CBS-control comparison, sensitivity varied across the different NODDI measures, with ICVF performing well across both brainstem and cortical regions in CBS and PSP-RS. In the comparison between PSP-RS and PD, the top-ranked ROIs mostly reflected those that showed most abnormalities in PSP-RS, likely since PSP-RS shows such widespread and severe white matter degeneration. There was still a suggestion that the PD participants showed greater involvement of AD-related regions with PD showing greater abnormalities in the fornix than PSP-RS. Prior studies comparing NODDI and FWE in PSP and PD (Mitchell, Archer et al. 2019) (Mitchell, Wilkes et al. 2022) highlighted free-water and extracellular volume markers in basal ganglia and midbrain as powerful differentiators; our results agree and further show that NODDI ICVF and IsoVF provide enhanced spatial coverage and capture clinically relevant midbrain-peduncular degeneration with finer specificity.

Differences were also observed between PSP-RS and CBS in the disease-specific regions, with PSP-RS showing greater involvement of SCP and midbrain, and CBS showing greater involvement of the corpus callosum and precentral and postcentral white matter. These findings concord with previous findings comparing PSP-RS and CBS using DTI (Erbetta, Mandelli et al. 2009, Whitwell, Schwarz et al. 2014). The top 5 ROIs with the highest cohen’s d that best differentiated CBS and PSP-RS were bilateral midbrain, caudate nucleus, medial lemniscus, SCP, and pons, detected by DTI MD, NODDI ICVF, and FBA FD counts, which show substantial correspondence with the brainstem and subcortical MRI atrophy patterns that distinguished CBS from PSP RS (Jabbari, Holland et al. 2020). Specifically, the midbrain, pons, and caudate (within their central structures ROI) map directly onto their highest value volumetric discriminators, while the SCP and medial lemniscus were not included in their atlas. Compared to the limited performance of CSF biomarkers for separating CBS from PSP RS (Giannakis, Konitsiotis et al. 2025), our dMRI tract findings highlight the potential clinical utility for improving differential diagnosis.

Structure-function analyses strengthened our interpretations. Across the cohort, cognitive performance was most strongly associated with frontal white matter integrity. Within CBS, ideomotor apraxia showed robust associations with NODDI ODI and FBA FD across precentral, cerebellar, and temporoparietal tracts. These findings suggest that increased axonal dispersion and reduced fiber density contribute to ideomotor apraxia. Differences between WAB-Praxis and TULIA correlations may stem from how each test probes praxis. WAB-Praxis relies on command-driven gesture production within a language framework, while TULIA emphasizes imitation and more complex gesture sequences. These differences likely tap into overlapping but not identical neural systems, especially in CBS where apraxia can arise from multiple cortical-subcortical disconnection pathways. Interpretation is further limited by the modest number of CBS participants with available scores (WAB-Praxis: 14/25; TULIA: 17/25). Across CBS, FBA FD counts and sums produced the clear and consistent associations with praxis severity, but FD mean was less informative. It is likely a result of spatial averaging dampening focal tract abnormalities. In PSP-RS, gait and postural instability correlated most strongly with NODDI IsoVF in the midbrain, basal ganglia, and thalamus, pointing to free-water-related degeneration of brainstem motor circuits as a driver of axial disability.

Methodologically, this study offers a unified framework for identifying the most discriminative structure-metric combinations across syndromes and for linking microstructure to multiple clinical scales. The large and well-characterized CBS and PSP-RS cohorts, combined with rigorous family-wise error correction, strengthen the reliability of the findings. Several limitations are worth mentioning. Some of our CBS patients (PSIS above 1) also had clinical features of PSP, particularly ocular motor impairment, as is common in CBS, but it may have reduced diffusion differences between CBS and PSP-RS in PSP-related regions. The cross-sectional design limits inferences about disease progression. Group imbalance, with more PSP-RS cases, may influence effect-size comparisons. ROI-based analyses may miss fine-grained tract abnormalities, and the absence of tau biomarkers restricts pathophysiological interpretation.

Collectively, these findings demonstrate that both conventional DTI and NODDI ICVF reliably separate CBS, PSP-RS, PD, and controls, but NODDI ICVF provides greater statistical sensitivity, i.e. yielding finer p-values while maintaining comparably large effect sizes. Importantly, structure-function analyses further supported the clinical relevance of these findings, linking syndrome-specific diffusion abnormalities to motor and cognitive impairments in CBS and PSP-RS. Together, these results indicate that neurite-density-based diffusion metrics may offer more precise and biologically informative imaging biomarkers for APS, with potential utility for earlier differential diagnosis and biomarker development.

## Acknowledgement

This study was funded by National Institutes of Health (NIH) grants R01-NS89757 and R01-AG87140.

## Notes

### Competing Interest Statement

The authors have declared no competing interest.

### Author Declarations

Institutional Review Board of Mayo Clinic gave ethical approval for this work.

