## Supplemental for "Multi-model Diffusion MRI Signatures in Atypical Parkinsonian Disorders"

### Supplemental Materials

#### Pairwise Comparison of Clinical Characteristics

| Variable | Comparison  Group2 vs Group1 | Ngroup1 | Ngroup2 | Cohen's d | Raw_p | Bonferroni_p | BH_p |
| --- | --- | --- | --- | --- | --- | --- | --- |
| Age at visit, years | CBS vs Control | 35/35 | 25/25 | -0.082 | 0.418 | 1.000 | 0.502 |
| Age at visit, years | PD vs Control | 35/35 | 21/21 | -0.335 | 0.115 | 0.693 | 0.231 |
| Age at visit, years | PSP-RS vs Control | 35/35 | 42/42 | -0.740 | 0.001 | 0.007 | 0.007 |
| Age at visit, years | PD vs CBS | 25/25 | 21/21 | -0.234 | 0.332 | 1.000 | 0.498 |
| Age at visit, years | PSP-RS vs CBS | 25/25 | 42/42 | -0.583 | 0.016 | 0.097 | 0.049 |
| Age at visit, years | PSP-RS vs PD | 21/21 | 42/42 | -0.232 | 0.535 | 1.000 | 0.535 |
| Age at onset, years | CBS vs Control | 0/35 | 25/25 |  |  |  |  |
| Age at onset, years | PD vs Control | 0/35 | 21/21 |  |  |  |  |
| Age at onset, years | PSP-RS vs Control | 0/35 | 42/42 |  |  |  |  |
| Age at onset, years | PD vs CBS | 25/25 | 21/21 | 0.169 | 0.691 | 1.000 | 1.000 |
| Age at onset, years | PSP-RS vs CBS | 25/25 | 42/42 | -0.514 | 0.037 | 0.225 | 0.112 |
| Age at onset, years | PSP-RS vs PD | 21/21 | 42/42 | -0.616 | 0.029 | 0.176 | 0.112 |
| Time from onset to visit, years | CBS vs Control | 0/35 | 25/25 |  |  |  |  |
| Time from onset to visit, years | PD vs Control | 0/35 | 21/21 |  |  |  |  |
| Time from onset to visit, years | PSP-RS vs Control | 0/35 | 42/42 |  |  |  |  |
| Time from onset to visit, years | PD vs CBS | 25/25 | 21/21 | -1.192 | 0.000 | 0.002 | 0.002 |
| Time from onset to visit, years | PSP-RS vs CBS | 25/25 | 42/42 | -0.346 | 0.268 | 1.000 | 0.535 |
| Time from onset to visit, years | PSP-RS vs PD | 21/21 | 42/42 | 1.052 | 0.001 | 0.005 | 0.002 |
| Education | CBS vs Control | 31/35 | 25/25 | 0.600 | 0.028 | 0.166 | 0.166 |
| Education | PD vs Control | 31/35 | 21/21 | 0.512 | 0.150 | 0.899 | 0.300 |
| Education | PSP-RS vs Control | 31/35 | 41/42 | 0.363 | 0.110 | 0.659 | 0.300 |
| Education | PD vs CBS | 25/25 | 21/21 | -0.077 | 0.692 | 1.000 | 0.817 |
| Education | PSP-RS vs CBS | 25/25 | 41/42 | -0.148 | 0.625 | 1.000 | 0.817 |
| Education | PSP-RS vs PD | 21/21 | 41/42 | -0.076 | 0.817 | 1.000 | 0.817 |
| MoCA | CBS vs Control | 35/35 | 25/25 | 1.670 | 0.000 | 0.000 | 0.000 |
| MoCA | PD vs Control | 35/35 | 21/21 | 0.951 | 0.005 | 0.030 | 0.009 |
| MoCA | PSP-RS vs Control | 35/35 | 42/42 | 1.717 | 0.000 | 0.000 | 0.000 |
| MoCA | PD vs CBS | 25/25 | 21/21 | -0.594 | 0.025 | 0.147 | 0.029 |
| MoCA | PSP-RS vs CBS | 25/25 | 42/42 | 0.098 | 0.745 | 1.000 | 0.745 |
| MoCA | PSP-RS vs PD | 21/21 | 42/42 | 0.713 | 0.006 | 0.035 | 0.009 |
| MDS-UPDRS III | CBS vs Control | 23/35 | 25/25 | -2.713 | 0.000 | 0.000 | 0.000 |
| MDS-UPDRS III | PD vs Control | 23/35 | 20/21 | -2.261 | 0.000 | 0.000 | 0.000 |
| MDS-UPDRS III | PSP-RS vs Control | 23/35 | 42/42 | -2.973 | 0.000 | 0.000 | 0.000 |
| MDS-UPDRS III | PD vs CBS | 25/25 | 20/21 | 0.418 | 0.126 | 0.754 | 0.151 |
| MDS-UPDRS III | PSP-RS vs CBS | 25/25 | 42/42 | -0.243 | 0.429 | 1.000 | 0.429 |
| MDS-UPDRS III | PSP-RS vs PD | 20/21 | 42/42 | -0.687 | 0.017 | 0.102 | 0.025 |
| PSP Rating Scale | CBS vs Control | 0/35 | 24/25 |  |  |  |  |
| PSP Rating Scale | PD vs Control | 0/35 | 20/21 |  |  |  |  |
| PSP Rating Scale | PSP-RS vs Control | 0/35 | 42/42 |  |  |  |  |
| PSP Rating Scale | PD vs CBS | 24/25 | 20/21 | 1.260 | 0.000 | 0.001 | 0.000 |
| PSP Rating Scale | PSP-RS vs CBS | 24/25 | 42/42 | -0.532 | 0.021 | 0.128 | 0.043 |
| PSP Rating Scale | PSP-RS vs PD | 20/21 | 42/42 | -2.105 | 0.000 | 0.000 | 0.000 |
| PSP Rating Scale gait/midline | CBS vs Control | 0/35 | 14/25 |  |  |  |  |
| PSP Rating Scale gait/midline | PD vs Control | 0/35 | 20/21 |  |  |  |  |
| PSP Rating Scale gait/midline | PSP-RS vs Control | 0/35 | 42/42 |  |  |  |  |
| PSP Rating Scale gait/midline | PD vs CBS | 14/25 | 20/21 | 1.261 | 0.002 | 0.011 | 0.006 |
| PSP Rating Scale gait/midline | PSP-RS vs CBS | 14/25 | 42/42 | -0.130 | 0.733 | 1.000 | 1.000 |
| PSP Rating Scale gait/midline | PSP-RS vs PD | 20/21 | 42/42 | -1.372 | 0.000 | 0.000 | 0.000 |
| FAB | CBS vs Control | 0/35 | 24/25 |  |  |  |  |
| FAB | PD vs Control | 0/35 | 19/21 |  |  |  |  |
| FAB | PSP-RS vs Control | 0/35 | 42/42 |  |  |  |  |
| FAB | PD vs CBS | 24/25 | 19/21 | -0.472 | 0.126 | 0.755 | 0.252 |
| FAB | PSP-RS vs CBS | 24/25 | 42/42 | 0.499 | 0.059 | 0.353 | 0.177 |
| FAB | PSP-RS vs PD | 19/21 | 42/42 | 1.054 | 0.000 | 0.001 | 0.001 |
| WAB_Praxis | CBS vs Control | 22/35 | 14/25 | 1.504 | 0.000 | 0.000 | 0.000 |
| WAB_Praxis | PD vs Control | 22/35 | 14/21 | -0.183 | 0.256 | 1.000 | 0.296 |
| WAB_Praxis | PSP-RS vs Control | 22/35 | 1/42 |  | 0.085 | 0.509 | 0.127 |
| WAB_Praxis | PD vs CBS | 14/25 | 14/21 | -1.340 | 0.000 | 0.000 | 0.000 |
| WAB_Praxis | PSP-RS vs CBS | 14/25 | 1/42 |  | 0.296 | 1.000 | 0.296 |
| WAB_Praxis | PSP-RS vs PD | 14/21 | 1/42 |  | 0.074 | 0.446 | 0.127 |
| PSIS | CBS vs Control | 22/35 | 23/25 | -1.310 | 0.000 | 0.001 | 0.000 |
| PSIS | PD vs Control | 22/35 | 20/21 | -0.871 | 0.009 | 0.053 | 0.011 |
| PSIS | PSP-RS vs Control | 22/35 | 41/42 | -4.356 | 0.000 | 0.000 | 0.000 |
| PSIS | PD vs CBS | 23/25 | 20/21 | 0.597 | 0.081 | 0.488 | 0.081 |
| PSIS | PSP-RS vs CBS | 23/25 | 41/42 | -2.520 | 0.000 | 0.000 | 0.000 |
| PSIS | PSP-RS vs PD | 20/21 | 41/42 | -3.401 | 0.000 | 0.000 | 0.000 |
| TULIA | CBS vs Control | 11/35 | 17/25 | 1.880 | 0.000 | 0.000 | 0.000 |
| TULIA | PD vs Control | 11/35 | 14/21 | -0.382 | 0.256 | 1.000 | 0.256 |
| TULIA | PSP-RS vs Control | 11/35 | 40/42 | 0.549 | 0.140 | 0.837 | 0.167 |
| TULIA | PD vs CBS | 17/25 | 14/21 | -2.070 | 0.000 | 0.000 | 0.000 |
| TULIA | PSP-RS vs CBS | 17/25 | 40/42 | -2.155 | 0.000 | 0.000 | 0.000 |
| TULIA | PSP-RS vs PD | 14/21 | 40/42 | 0.770 | 0.009 | 0.056 | 0.014 |
| TULIA_left | CBS vs Control | 11/35 | 16/25 | 1.217 | 0.001 | 0.005 | 0.002 |
| TULIA_left | PD vs Control | 11/35 | 14/21 | -0.382 | 0.256 | 1.000 | 0.256 |
| TULIA_left | PSP-RS vs Control | 11/35 | 39/42 | 0.456 | 0.245 | 1.000 | 0.256 |
| TULIA_left | PD vs CBS | 16/25 | 14/21 | -1.372 | 0.000 | 0.000 | 0.000 |
| TULIA_left | PSP-RS vs CBS | 16/25 | 39/42 | -1.352 | 0.000 | 0.002 | 0.001 |
| TULIA_left | PSP-RS vs PD | 14/21 | 39/42 | 0.679 | 0.019 | 0.113 | 0.028 |
| TULIA_right | CBS vs Control | 11/35 | 16/25 | 1.131 | 0.012 | 0.070 | 0.023 |
| TULIA_right | PD vs Control | 11/35 | 14/21 | -0.382 | 0.256 | 1.000 | 0.292 |
| TULIA_right | PSP-RS vs Control | 11/35 | 39/42 | 0.438 | 0.292 | 1.000 | 0.292 |
| TULIA_right | PD vs CBS | 16/25 | 14/21 | -1.287 | 0.001 | 0.005 | 0.005 |
| TULIA_right | PSP-RS vs CBS | 16/25 | 39/42 | -1.219 | 0.010 | 0.058 | 0.023 |
| TULIA_right | PSP-RS vs PD | 14/21 | 39/42 | 0.662 | 0.025 | 0.151 | 0.038 |

**Table S1** **Pairwise post hoc comparisons for continuous demographic and clinical variables.** This table reports pairwise group comparisons following the omnibus Kruskal–Wallis tests in Table 1. For each continuous variable, Mann–Whitney U tests were performed for all group pairs, and effect sizes were quantified using Cohen’s d. Ngroup1 and Ngroup2 indicate sample sizes for each side of the comparison, expressed as non-NaN entries / total participants in that group. Multiple-comparison corrections were applied per variable across all group pairs using both Bonferroni correction and the Benjamini–Hochberg (BH) false discovery rate method. Columns include the uncorrected p-value (Raw_p), Bonferroni-adjusted p-value (Bonferroni_p), and Benjamini–Hochberg (BH)-adjusted p-value (BH_p).

| **Variable** | **Comparison** | **Ngroup1** | **Ngroup2** | **Test** | **Raw_p** | **BH_p** |
| --- | --- | --- | --- | --- | --- | --- |
| Gender | CBS vs Control | 35/35 | 25/25 | Fisher | 0.602 | 0.723 |
| Gender | PD vs Control | 35/35 | 21/21 | Fisher | 0.053 | 0.106 |
| Gender | PSP-RS vs Control | 35/35 | 42/42 | Fisher | 0.102 | 0.153 |
| Gender | PD vs CBS | 25/25 | 21/21 | Fisher | 0.019 | 0.072 |
| Gender | PSP-RS vs CBS | 25/25 | 42/42 | Fisher | 0.024 | 0.072 |
| Gender | PSP-RS vs PD | 21/21 | 42/42 | Fisher | 0.768 | 0.768 |
| Handedness | CBS vs Control | 28/35 | 22/25 | Chi-square | 0.159 | 0.191 |
| Handedness | PSP-RS vs Control | 28/35 | 40/42 | Fisher | 1.000 | 1.000 |
| Handedness | PSP-RS vs CBS | 22/25 | 40/42 | Chi-square | 0.097 | 0.191 |
| Family history | PSP-RS vs CBS | 18/25 | 30/42 | Fisher | 0.555 | 1.000 |

**Table S2** **Pairwise categorical post hoc tests.** This table reports pairwise comparisons between diagnostic groups using χ² tests or Fisher’s exact tests, depending on expected cell counts. For each pair of groups, Ngroup1 and Ngroup2 denote the number of non-missing observations out of the total number of participants in that group (**non-NaN / total**), indicating how many participants contributed data to each comparison. Raw p-values and BH–corrected p-values are provided for each variable.

Because categorical tests are unreliable when either comparison group has very small usable sample size, pairwise contrasts in which either group had fewer than five non-missing observations were excluded from the table.

#### Top ROIs x Metrics for PD vs Control

Figures S7 showed Top ROIs x Metrics for PD vs Control.

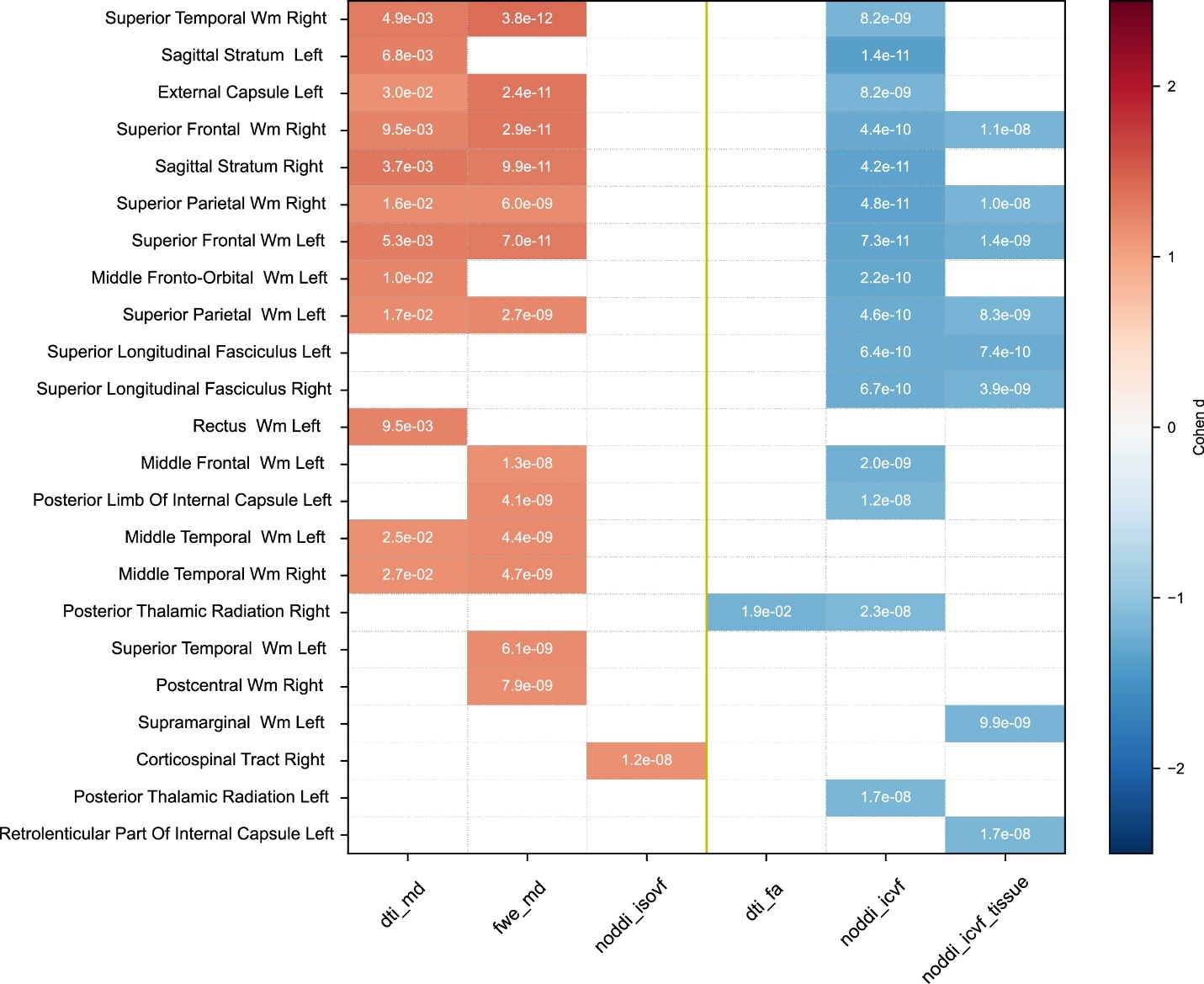

#### Figure S1 Top 50 ROI–metric pairs ranked by |Cohen’s d| for PD vs controls. Cell colors represent the magnitude of the effect size (|Cohen’s d|), and each cell displays pval_fwe. Metrics are arranged by polarity, with the left of the yellow midline indicating metrics for which higher values reflect worse brain integrity relative to controls.

#### Boxplots in key ROIs

In Supplemental Figures S2–S7, boxplots for ROIs bilateral precentral WM, SCP and midbrain are shown across all dMRI metrics, with FDR-corrected *q* values displayed as letter codes.

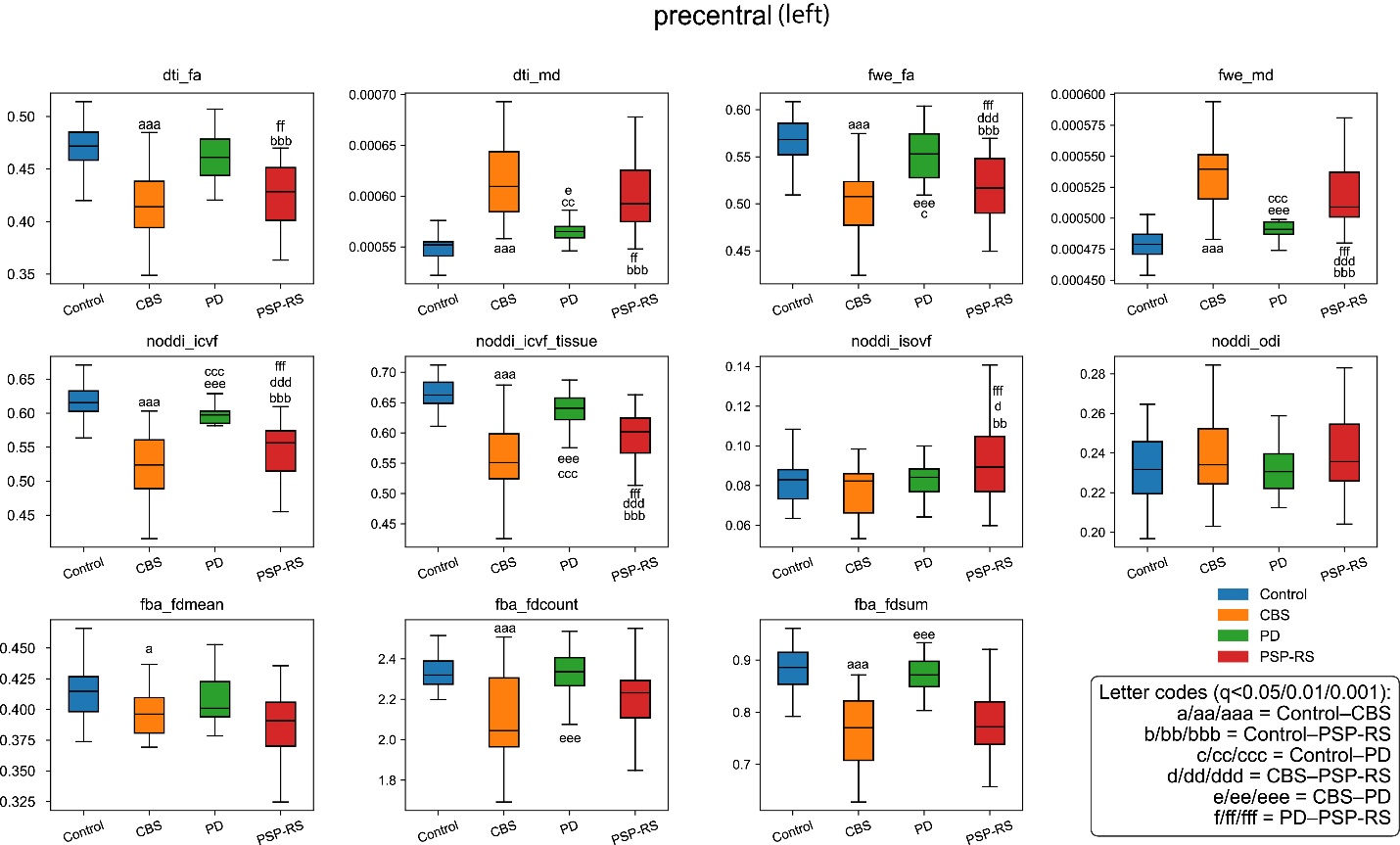

**Figure S2** Boxplots for each metric value of ROI Left Precentral WM.

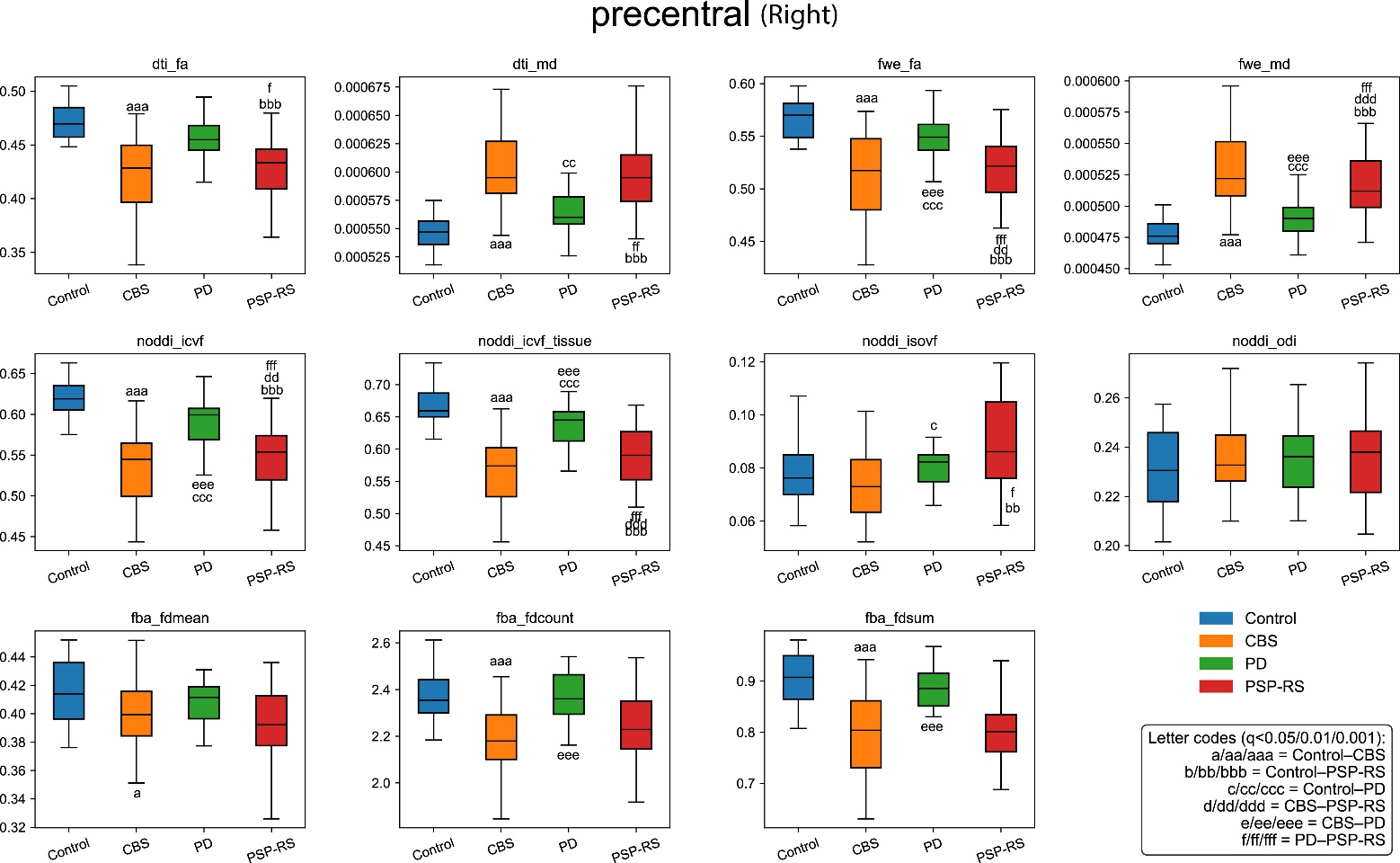

**Figure S3** Boxplots for each metric value of ROI Right Precentral WM.

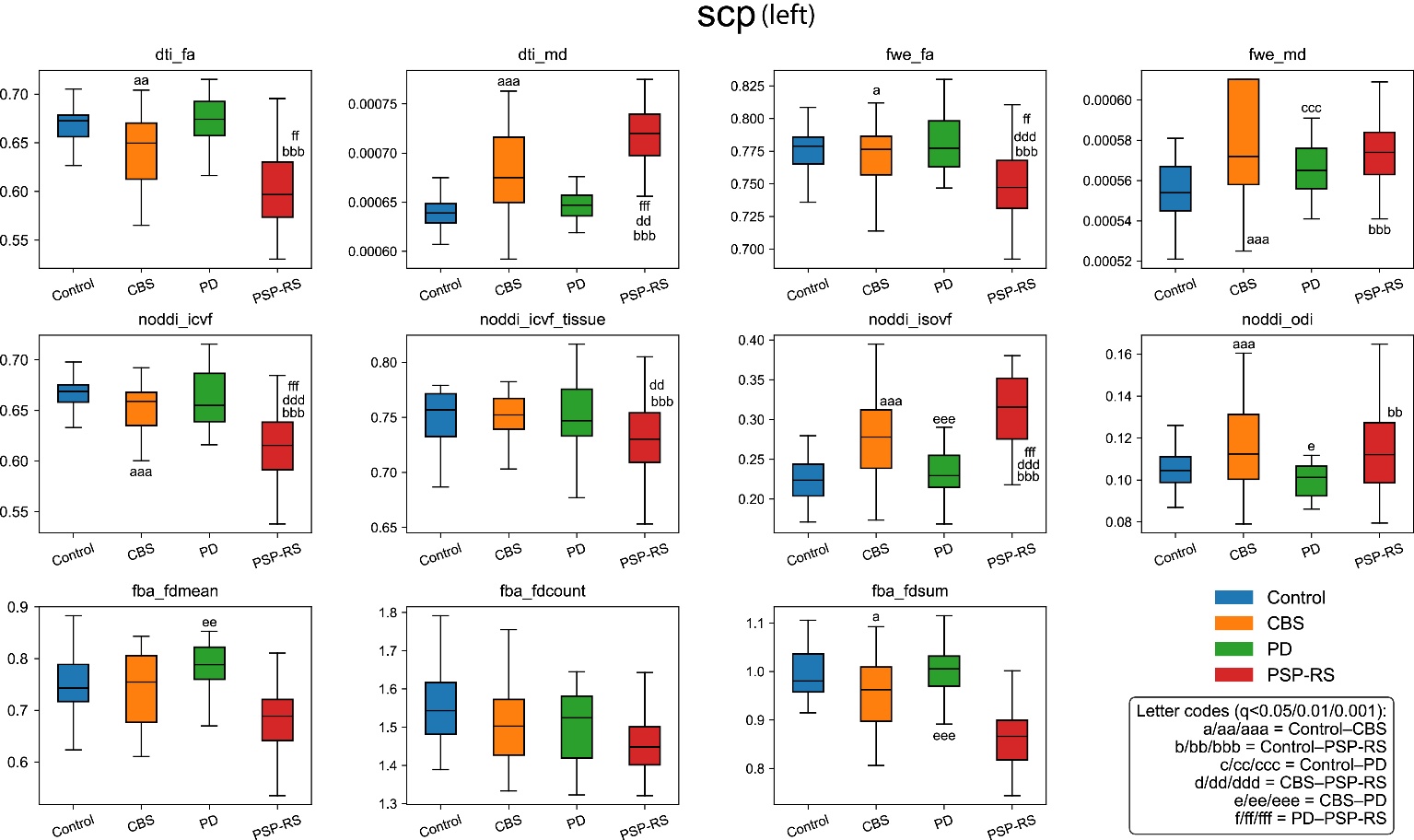

**Figure S4** Boxplots for each metric value of ROI Left SCP.

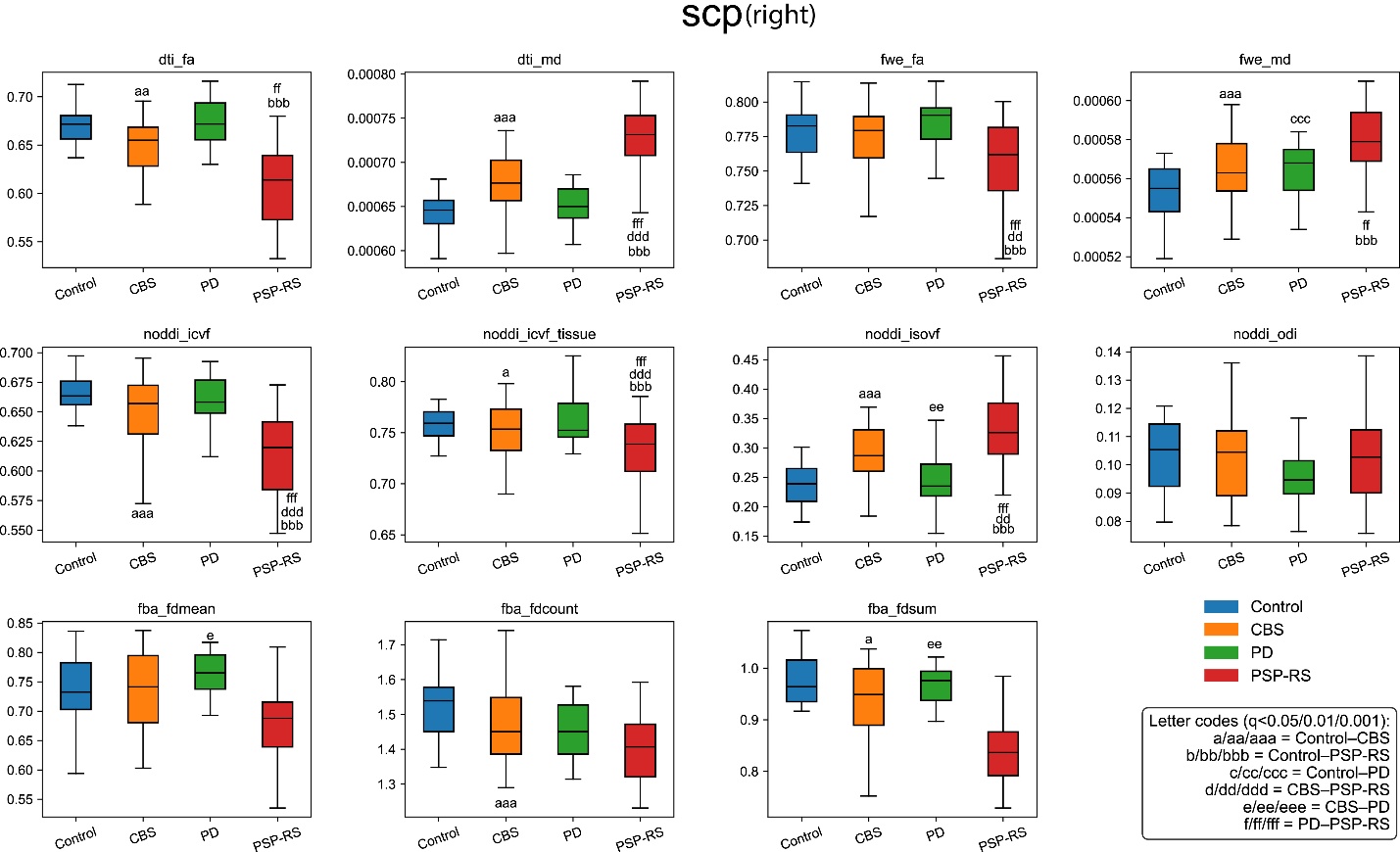

**Figure S5** Boxplots for each metric value of ROI Right SCP.

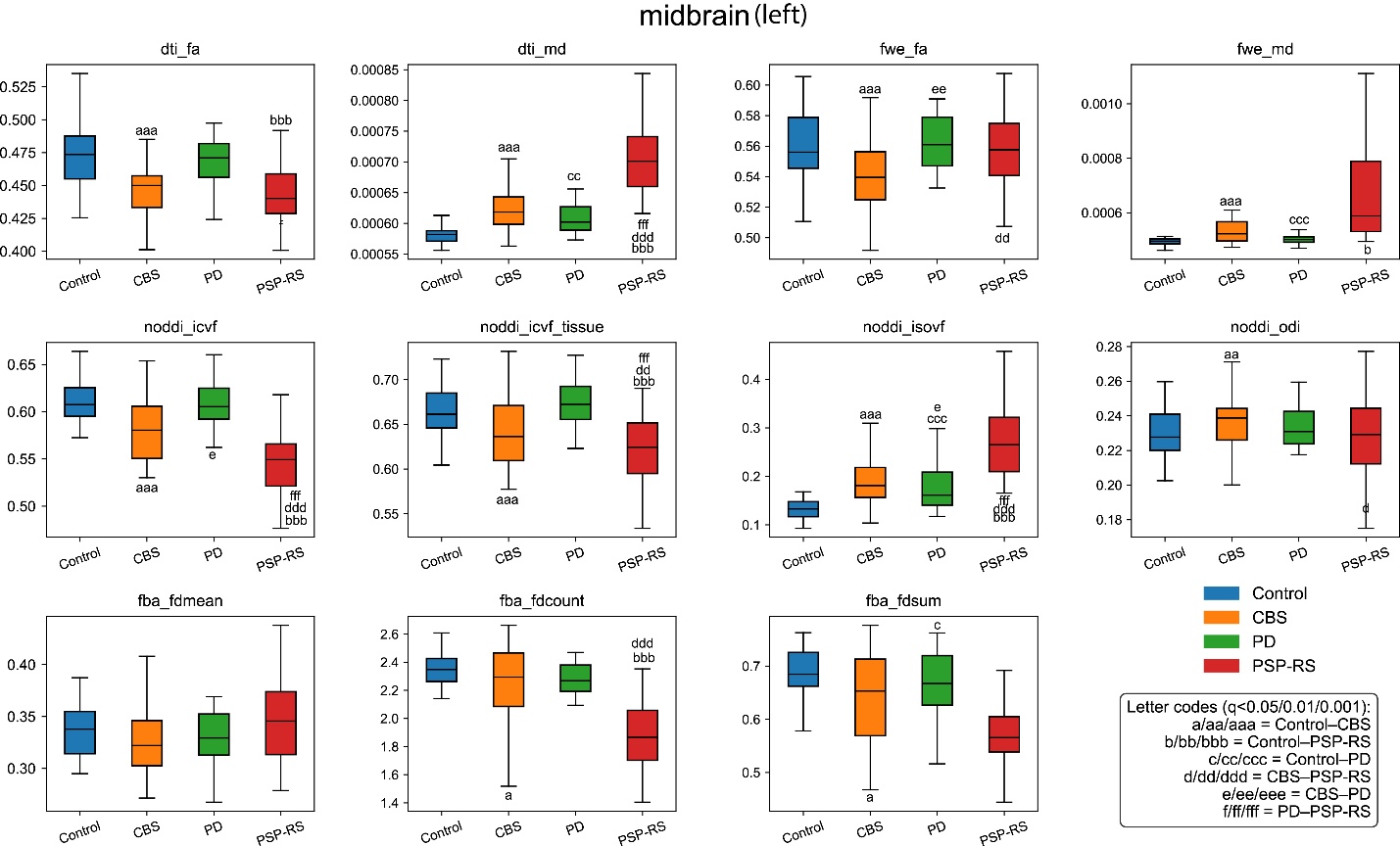

**Figure S6** Boxplots for each metric value of ROI Left Midbrain.

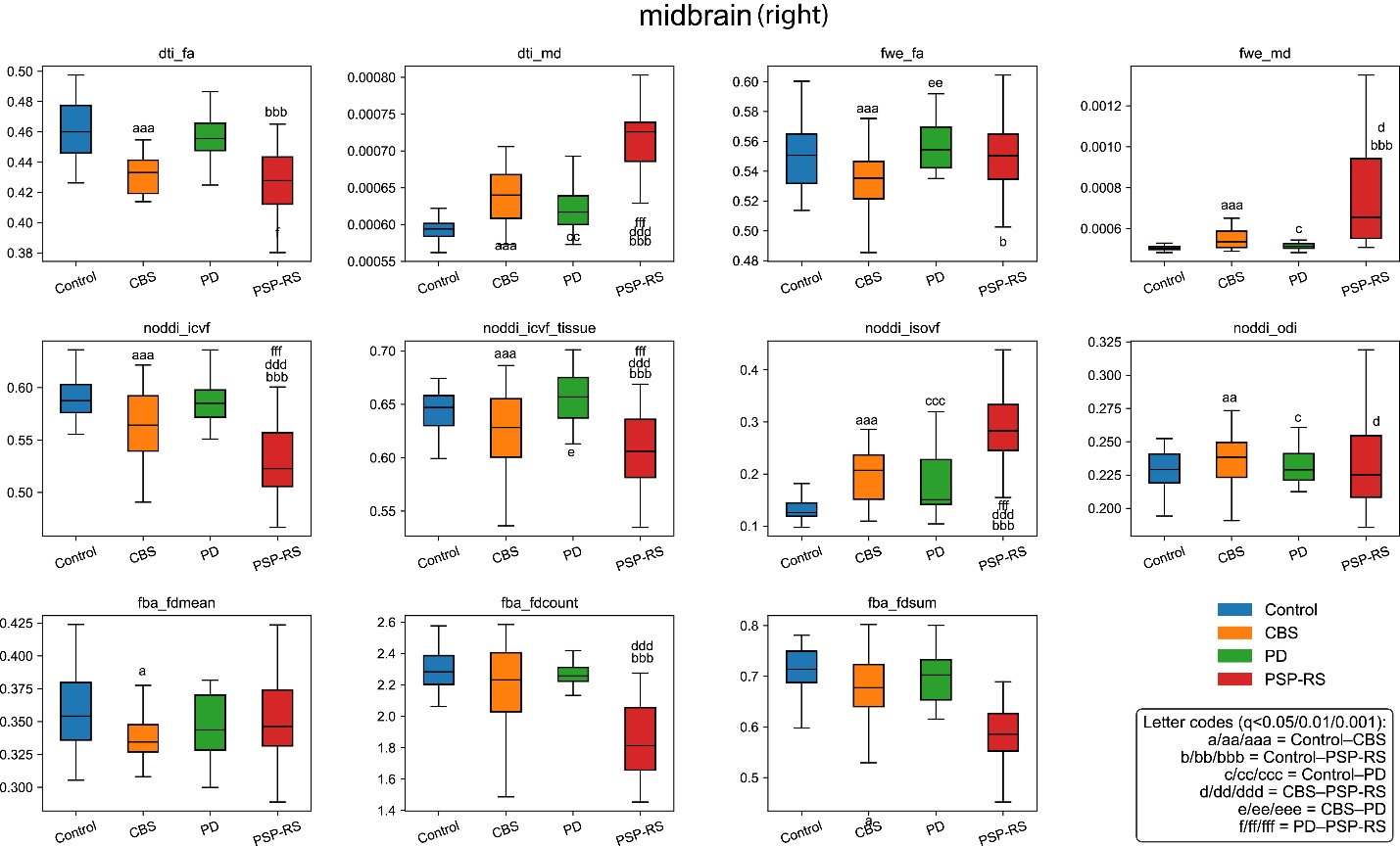

**Figure S7** Boxplots for each metric value of ROI Right Midbrain.

#### ANCOVA top50 Pairwise Tables

| group_a | group_b | adjusted_mean_diff | cohen_d | roi_id | roi_name | metric | qval_bh | pval_fwe |
| --- | --- | --- | --- | --- | --- | --- | --- | --- |
| CBS | Control | -0.09678 | -2.60331 | 6 | PrCWM_L PRECENTRAL__WM_left | noddi_icvf_mean | 3.53E-36 | 3.53E-36 |
| CBS | Control | -0.11132 | -2.53825 | 6 | PrCWM_L PRECENTRAL__WM_left | noddi_icvf_tissue_mean | 5.26E-35 | 5.26E-35 |
| CBS | Control | -0.09037 | -2.4891 | 97 | SCR_R Superior_corona_radiata_right | noddi_icvf_tissue_mean | 2.04E-34 | 4.09E-34 |
| CBS | Control | 5.84E-05 | 2.484339 | 6 | PrCWM_L PRECENTRAL__WM_left | fwe_md_mean | 4.99E-34 | 4.99E-34 |
| CBS | Control | -0.06188 | -2.44137 | 94 | PLIC_R Posterior_limb_of_internal_capsule_right | noddi_icvf_tissue_mean | 1.01E-33 | 3.02E-33 |
| CBS | Control | -0.09723 | -2.42409 | 68 | PrCWM_R PRECENTRAL_WM_right | noddi_icvf_tissue_mean | 1.56E-33 | 6.24E-33 |
| CBS | Control | -0.08558 | -2.39785 | 37 | SCR_L Superior_corona_radiata_left | noddi_icvf_mean | 8.55E-33 | 1.88E-32 |
| CBS | Control | -0.08501 | -2.38882 | 68 | PrCWM_R PRECENTRAL_WM_right | noddi_icvf_mean | 8.55E-33 | 2.75E-32 |
| CBS | Control | -0.05107 | -2.38237 | 94 | PLIC_R Posterior_limb_of_internal_capsule_right | noddi_icvf_mean | 8.55E-33 | 3.61E-32 |
| CBS | Control | -0.07904 | -2.37835 | 97 | SCR_R Superior_corona_radiata_right | noddi_icvf_mean | 8.55E-33 | 4.28E-32 |
| CBS | Control | 6.63E-05 | 2.350897 | 6 | PrCWM_L PRECENTRAL__WM_left | dti_md_mean | 5.82E-10 | 5.82E-10 |
| CBS | Control | -0.05772 | -2.33516 | 34 | PLIC_L Posterior_limb_of_internal_capsule_left | noddi_icvf_mean | 4.41E-32 | 2.65E-31 |
| CBS | Control | -0.09571 | -2.32199 | 37 | SCR_L Superior_corona_radiata_left | noddi_icvf_tissue_mean | 9.25E-32 | 4.62E-31 |
| CBS | Control | 5.09E-05 | 2.283555 | 68 | PrCWM_R PRECENTRAL_WM_right | fwe_md_mean | 1.18E-30 | 2.36E-30 |
| CBS | Control | 3.72E-05 | 2.276006 | 34 | PLIC_L Posterior_limb_of_internal_capsule_left | dti_md_mean | 8.20E-10 | 1.64E-09 |
| CBS | Control | -0.05643 | -2.16615 | 3 | SFWM_L SUPERIOR_FRONTAL_WM_left | noddi_icvf_mean | 4.96E-29 | 3.47E-28 |
| CBS | Control | 3.83E-05 | 2.160738 | 34 | PLIC_L Posterior_limb_of_internal_capsule_left | fwe_md_mean | 1.46E-28 | 4.37E-28 |
| CBS | Control | -0.06859 | -2.1558 | 34 | PLIC_L Posterior_limb_of_internal_capsule_left | noddi_icvf_tissue_mean | 8.99E-29 | 5.39E-28 |
| CBS | Control | -0.07311 | -2.13883 | 112 | BCC_R Body_of_corpus_callosum_right | noddi_icvf_mean | 1.39E-28 | 1.11E-27 |
| CBS | Control | -0.05158 | -2.12511 | 3 | SFWM_L SUPERIOR_FRONTAL_WM_left | fwe_fa_mean | 2.00E-27 | 2.00E-27 |
| CBS | Control | -0.06611 | -2.10551 | 3 | SFWM_L SUPERIOR_FRONTAL_WM_left | noddi_icvf_tissue_mean | 6.59E-28 | 4.61E-27 |
| CBS | Control | -0.09449 | -2.10109 | 52 | BCC_L Body_of_corpus_callosum_left | dti_fa_mean | 1.11E-08 | 1.89E-08 |
| CBS | Control | -0.09871 | -2.08962 | 112 | BCC_R Body_of_corpus_callosum_right | dti_fa_mean | 1.11E-08 | 2.22E-08 |
| CBS | Control | -0.06491 | -2.08693 | 6 | PrCWM_L PRECENTRAL__WM_left | fwe_fa_mean | 5.10E-27 | 1.02E-26 |
| CBS | Control | -0.05471 | -2.0726 | 65 | SFWM_R SUPERIOR_FRONTAL__WM_right | noddi_icvf_mean | 2.09E-27 | 1.88E-26 |
| CBS | Control | 3.53E-05 | 2.062925 | 94 | PLIC_R Posterior_limb_of_internal_capsule_right | fwe_md_mean | 7.11E-27 | 2.84E-26 |
| CBS | Control | -0.07163 | -2.04484 | 52 | BCC_L Body_of_corpus_callosum_left | noddi_icvf_mean | 6.16E-27 | 6.16E-26 |
| CBS | Control | -0.07531 | -2.03708 | 42 | SLF_L Superior_longitudinal_fasciculus_left | noddi_icvf_mean | 7.80E-27 | 8.58E-26 |
| CBS | Control | -0.07177 | -2.03234 | 7 | PoCWM_L POSTCENTRAL__WM_left | noddi_icvf_mean | 8.76E-27 | 1.05E-25 |
| CBS | Control | 3.79E-05 | 2.030817 | 3 | SFWM_L SUPERIOR_FRONTAL_WM_left | dti_md_mean | 1.69E-08 | 5.06E-08 |
| CBS | Control | -0.06385 | -2.02935 | 65 | SFWM_R SUPERIOR_FRONTAL__WM_right | noddi_icvf_tissue_mean | 1.49E-26 | 1.19E-25 |
| CBS | Control | 0.06347 | 2.021184 | 116 | Midbrain_R MIDBRAIN_right | noddi_isovf_mean | 1.69E-25 | 1.69E-25 |
| CBS | Control | -0.06136 | -2.01272 | 4 | MFWM_L MIDDLE_FRONTAL__WM_left | noddi_icvf_mean | 1.87E-26 | 2.43E-25 |
| CBS | Control | 3.26E-05 | 2.00154 | 3 | SFWM_L SUPERIOR_FRONTAL_WM_left | fwe_md_mean | 6.59E-26 | 3.92E-25 |
| CBS | Control | 4.60E-05 | 2.001344 | 7 | PoCWM_L POSTCENTRAL__WM_left | fwe_md_mean | 6.59E-26 | 3.95E-25 |
| CBS | Control | -0.08041 | -1.98004 | 7 | PoCWM_L POSTCENTRAL__WM_left | noddi_icvf_tissue_mean | 1.09E-25 | 9.82E-25 |
| CBS | Control | -0.07134 | -1.97254 | 102 | SLF_R Superior_longitudinal_fasciculus_right | noddi_icvf_mean | 9.67E-26 | 1.35E-24 |
| CBS | Control | -0.08864 | -1.96853 | 42 | SLF_L Superior_longitudinal_fasciculus_left | noddi_icvf_tissue_mean | 1.56E-25 | 1.61E-24 |
| CBS | Control | -0.05627 | -1.96742 | 6 | PrCWM_L PRECENTRAL__WM_left | dti_fa_mean | 4.12E-08 | 1.23E-07 |
| CBS | Control | -0.08517 | -1.96701 | 23 | SMWM_L SUPRAMARGINAL__WM_left | noddi_icvf_tissue_mean | 1.56E-25 | 1.71E-24 |
| CBS | Control | 3.49E-05 | 1.958591 | 4 | MFWM_L MIDDLE_FRONTAL__WM_left | fwe_md_mean | 3.51E-25 | 2.46E-24 |
| CBS | Control | 5.83E-05 | 1.95214 | 68 | PrCWM_R PRECENTRAL_WM_right | dti_md_mean | 3.83E-08 | 1.53E-07 |
| CBS | Control | -0.05186 | -1.94639 | 7 | PoCWM_L POSTCENTRAL__WM_left | fwe_fa_mean | 1.38E-24 | 4.14E-24 |
| CBS | Control | -0.05629 | -1.9315 | 68 | PrCWM_R PRECENTRAL_WM_right | fwe_fa_mean | 1.95E-24 | 7.81E-24 |
| CBS | Control | 3.54E-05 | 1.924585 | 94 | PLIC_R Posterior_limb_of_internal_capsule_right | dti_md_mean | 4.45E-08 | 2.26E-07 |
| CBS | Control | 5.31E-05 | 1.912641 | 7 | PoCWM_L POSTCENTRAL__WM_left | dti_md_mean | 4.45E-08 | 2.67E-07 |
| CBS | Control | -0.06524 | -1.91045 | 39 | CGC_L Cingulum_(cingulate_gyrus)_left | fwe_fa_mean | 3.84E-24 | 1.92E-23 |
| CBS | Control | -0.08313 | -1.90671 | 102 | SLF_R Superior_longitudinal_fasciculus_right | noddi_icvf_tissue_mean | 1.88E-24 | 2.25E-23 |
| CBS | Control | 5.25E-05 | 1.902821 | 97 | SCR_R Superior_corona_radiata_right | fwe_md_mean | 3.32E-24 | 2.66E-23 |
| CBS | Control | -0.04424 | -1.9009 | 3 | SFWM_L SUPERIOR_FRONTAL_WM_left | dti_fa_mean | 7.87E-08 | 3.15E-07 |

**Table S3** CBS vs Control

| group_a | group_b | adjusted_mean_diff | cohen_d | roi_id | roi_name | metric | qval_bh | pval_fwe |
| --- | --- | --- | --- | --- | --- | --- | --- | --- |
| CBS | PD | -0.1072 | -1.90023 | 6 | PrCWM_L PRECENTRAL__WM_left | fba_fdsum_mean | 3.46E-10 | 3.46E-10 |
| CBS | PD | -0.19481 | -1.87924 | 88 | CST_R Corticospinal_tract_right | fba_fdcount_mean | 5.40E-10 | 5.40E-10 |
| CBS | PD | -0.08606 | -1.83483 | 112 | BCC_R Body_of_corpus_callosum_right | fba_fdmean_mean | 1.37E-09 | 1.37E-09 |
| CBS | PD | -0.07825 | -1.69732 | 112 | BCC_R Body_of_corpus_callosum_right | fba_fdsum_mean | 1.19E-08 | 2.38E-08 |
| CBS | PD | -0.07006 | -1.69618 | 6 | PrCWM_L PRECENTRAL__WM_left | noddi_icvf_mean | 4.23E-10 | 4.23E-10 |
| CBS | PD | -0.08064 | -1.6387 | 6 | PrCWM_L PRECENTRAL__WM_left | noddi_icvf_tissue_mean | 1.69E-09 | 1.69E-09 |
| CBS | PD | 4.46E-05 | 1.635587 | 6 | PrCWM_L PRECENTRAL__WM_left | fwe_md_mean | 1.82E-09 | 1.82E-09 |
| CBS | PD | 0.532738 | 1.576163 | 2 | Caudate_L CAUDATE_NUCLEUS_left | fba_fdcount_mean | 1.37E-07 | 2.75E-07 |
| CBS | PD | 4.97E-05 | 1.542377 | 6 | PrCWM_L PRECENTRAL__WM_left | dti_md_mean | 0.02899 | 0.02899 |
| CBS | PD | -0.06892 | -1.51488 | 68 | PrCWM_R PRECENTRAL_WM_right | noddi_icvf_tissue_mean | 1.57E-08 | 3.14E-08 |
| CBS | PD | 0.324584 | 1.505013 | 40 | CGH_L Cingulum_(hippocampus)_left | fba_fdcount_mean | 3.72E-07 | 1.12E-06 |
| CBS | PD | -0.05995 | -1.48284 | 68 | PrCWM_R PRECENTRAL_WM_right | noddi_icvf_mean | 3.30E-08 | 6.59E-08 |
| CBS | PD | -0.0567 | -1.48099 | 22 | MFOWM_L MIDDLE_FRONTO-ORBITAL__WM_left | fba_fdmean_mean | 8.89E-07 | 1.78E-06 |
| CBS | PD | 3.79E-05 | 1.468485 | 68 | PrCWM_R PRECENTRAL_WM_right | fwe_md_mean | 4.58E-08 | 9.16E-08 |
| CBS | PD | 0.104877 | 1.452133 | 78 | ENT_R ENTORHINAL_AREA_right | fba_fdsum_mean | 1.03E-06 | 3.10E-06 |
| CBS | PD | -0.04946 | -1.45176 | 6 | PrCWM_L PRECENTRAL__WM_left | fwe_fa_mean | 1.34E-07 | 1.34E-07 |
| CBS | PD | 0.118798 | 1.423604 | 86 | Amygd_R AMYGDALA_right | fba_fdsum_mean | 1.34E-06 | 5.35E-06 |
| CBS | PD | 0.2257 | 1.422432 | 80 | ITWM_R INFERIOR_TEMPORAL_WM_right | fba_fdcount_mean | 1.14E-06 | 5.47E-06 |
| CBS | PD | -0.25075 | -1.42016 | 6 | PrCWM_L PRECENTRAL__WM_left | fba_fdcount_mean | 1.14E-06 | 5.71E-06 |
| CBS | PD | -0.07095 | -1.41283 | 52 | BCC_L Body_of_corpus_callosum_left | fba_fdmean_mean | 2.19E-06 | 6.56E-06 |
| CBS | PD | -0.0981 | -1.40562 | 62 | Pons_L PONS_left | fba_fdsum_mean | 1.43E-06 | 7.52E-06 |
| CBS | PD | -0.06585 | -1.39855 | 7 | PoCWM_L POSTCENTRAL__WM_left | fba_fdsum_mean | 1.43E-06 | 8.60E-06 |
| CBS | PD | -0.0163 | -1.37039 | 87 | Hippo_R HIPPOCAMPUS_right | fba_fdmean_mean | 3.64E-06 | 1.46E-05 |
| CBS | PD | 0.222582 | 1.368842 | 100 | CGH_R Cingulum_(hippocampus)_right | fba_fdcount_mean | 2.13E-06 | 1.50E-05 |
| CBS | PD | 0.120319 | 1.368376 | 101 | Fx_ST_R Fornix(cres)_Stria_terminalis__right | fba_fdcount_mean | 2.13E-06 | 1.51E-05 |
| CBS | PD | 0.354747 | 1.362107 | 87 | Hippo_R HIPPOCAMPUS_right | fba_fdcount_mean | 2.13E-06 | 1.70E-05 |
| CBS | PD | -0.09053 | -1.34299 | 68 | PrCWM_R PRECENTRAL_WM_right | fba_fdsum_mean | 3.46E-06 | 2.42E-05 |
| CBS | PD | -0.20008 | -1.33768 | 68 | PrCWM_R PRECENTRAL_WM_right | fba_fdcount_mean | 2.97E-06 | 2.67E-05 |
| CBS | PD | -0.03189 | -1.28214 | 3 | SFWM_L SUPERIOR_FRONTAL_WM_left | fwe_fa_mean | 2.81E-06 | 5.72E-06 |
| CBS | PD | -0.06851 | -1.27927 | 52 | BCC_L Body_of_corpus_callosum_left | fba_fdsum_mean | 9.68E-06 | 7.75E-05 |
| CBS | PD | -0.06345 | -1.26397 | 103 | SFO_R Superior_fronto-occipital_fasciculus__right | fwe_fa_mean | 2.81E-06 | 8.42E-06 |
| CBS | PD | 0.070478 | 1.255896 | 2 | Caudate_L CAUDATE_NUCLEUS_left | fba_fdsum_mean | 1.31E-05 | 0.000118 |
| CBS | PD | 0.048483 | 1.24498 | 43 | SFO_L Superior_fronto-occipital_fasciculus_left | noddi_odi_mean | 1.26E-05 | 1.26E-05 |
| CBS | PD | 0.366062 | 1.243482 | 78 | ENT_R ENTORHINAL_AREA_right | fba_fdcount_mean | 1.47E-05 | 0.000147 |
| CBS | PD | 0.028384 | 1.242926 | 23 | SMWM_L SUPRAMARGINAL__WM_left | fba_fdmean_mean | 2.96E-05 | 0.000148 |
| CBS | PD | -0.02148 | -1.22844 | 9 | PreCuWM_L PRE-CUNEUS__WM_left | noddi_isovf_mean | 1.78E-05 | 1.78E-05 |
| CBS | PD | -0.03849 | -1.21971 | 94 | PLIC_R Posterior_limb_of_internal_capsule_right | noddi_icvf_tissue_mean | 7.12E-06 | 2.14E-05 |
| CBS | PD | -0.03743 | -1.21188 | 7 | PoCWM_L POSTCENTRAL__WM_left | fwe_fa_mean | 6.29E-06 | 2.51E-05 |
| CBS | PD | 0.138685 | 1.205503 | 39 | CGC_L Cingulum_(cingulate_gyrus)_left | fba_fdcount_mean | 2.60E-05 | 0.000286 |
| CBS | PD | -0.03499 | -1.19405 | 52 | BCC_L Body_of_corpus_callosum_left | fwe_fa_mean | 7.27E-06 | 3.64E-05 |
| CBS | PD | -0.05123 | -1.1861 | 7 | PoCWM_L POSTCENTRAL__WM_left | noddi_icvf_tissue_mean | 1.07E-05 | 4.28E-05 |
| CBS | PD | 0.121192 | 1.183819 | 99 | CGC_R Cingulum_(cingulate_gyrus)_right | fba_fdcount_mean | 3.47E-05 | 0.000416 |
| CBS | PD | -0.045 | -1.1779 | 7 | PoCWM_L POSTCENTRAL__WM_left | noddi_icvf_mean | 1.69E-05 | 5.06E-05 |
| CBS | PD | 0.039387 | 1.171524 | 103 | SFO_R Superior_fronto-occipital_fasciculus__right | noddi_odi_mean | 2.88E-05 | 5.76E-05 |
| CBS | PD | -0.04206 | -1.16486 | 34 | PLIC_L Posterior_limb_of_internal_capsule_left | noddi_icvf_tissue_mean | 1.32E-05 | 6.60E-05 |
| CBS | PD | -0.03748 | -1.14112 | 68 | PrCWM_R PRECENTRAL_WM_right | fwe_fa_mean | 1.77E-05 | 0.000106 |
| CBS | PD | 0.418658 | 1.124637 | 83 | MFOWM_R MIDDLE_FRONTO-ORBITAL_WM_right | fba_fdcount_mean | 8.75E-05 | 0.001137 |
| CBS | PD | 2.92E-05 | 1.121124 | 7 | PoCWM_L POSTCENTRAL__WM_left | fwe_md_mean | 5.29E-05 | 0.000159 |
| CBS | PD | -0.03427 | -1.10828 | 112 | BCC_R Body_of_corpus_callosum_right | fwe_fa_mean | 2.92E-05 | 0.000204 |
| CBS | PD | 0.09103 | 1.102553 | 41 | Fx_ST_L Fornix(cres)_Stria_terminalisleft | fba_fdcount_mean | 0.000117 | 0.001642 |

**Table S4** CBS vs PD

| group_a | group_b | adjusted_mean_diff | cohen_d | roi_id | roi_name | metric | qval_bh | pval_fwe |
| --- | --- | --- | --- | --- | --- | --- | --- | --- |
| CBS | PSP-RS | -7.38E-05 | -1.62537 | 116 | Midbrain_R MIDBRAIN_right | dti_md_mean | 2.52E-05 | 2.52E-05 |
| CBS | PSP-RS | -6.68E-05 | -1.38671 | 61 | Midbrain_L MIDBRAIN_left | dti_md_mean | 0.000319 | 0.000638 |
| CBS | PSP-RS | 0.041923 | 1.374108 | 116 | Midbrain_R MIDBRAIN_right | noddi_icvf_mean | 4.20E-13 | 4.20E-13 |
| CBS | PSP-RS | 0.48629 | 1.361731 | 13 | Caudate_R CAUDATE_NUCLEUS_right | fba_fdcount_mean | 4.01E-12 | 5.78E-12 |
| CBS | PSP-RS | 0.5349 | 1.352695 | 58 | GlobusPalidus_R GLOBUS_PALLIDUS_right | fba_fdcount_mean | 4.01E-12 | 8.02E-12 |
| CBS | PSP-RS | -2.40E-05 | -1.3326 | 90 | ML_R Medial_lemniscus_right | dti_md_mean | 0.00037 | 0.001297 |
| CBS | PSP-RS | 0.423244 | 1.322901 | 2 | Caudate_L CAUDATE_NUCLEUS_left | fba_fdcount_mean | 7.84E-12 | 2.35E-11 |
| CBS | PSP-RS | -4.50E-05 | -1.32236 | 91 | SCP_R Superior_cerebellar_peduncle_right | dti_md_mean | 0.00037 | 0.001481 |
| CBS | PSP-RS | -0.07171 | -1.25816 | 116 | Midbrain_R MIDBRAIN_right | noddi_isovf_mean | 3.61E-11 | 3.61E-11 |
| CBS | PSP-RS | -2.91E-05 | -1.17851 | 117 | Pons_R PONS_right | dti_md_mean | 0.001826 | 0.009169 |
| CBS | PSP-RS | -5.59E-05 | -1.16406 | 58 | GlobusPalidus_R GLOBUS_PALLIDUS_right | dti_md_mean | 0.001826 | 0.010956 |
| CBS | PSP-RS | 0.033961 | 1.104425 | 31 | SCP_L Superior_cerebellar_peduncle_left | noddi_icvf_mean | 3.76E-09 | 9.70E-09 |
| CBS | PSP-RS | 0.036989 | 1.100136 | 91 | SCP_R Superior_cerebellar_peduncle_right | noddi_icvf_mean | 3.76E-09 | 1.13E-08 |
| CBS | PSP-RS | 0.036082 | 1.0795 | 61 | Midbrain_L MIDBRAIN_left | noddi_icvf_mean | 5.78E-09 | 2.31E-08 |
| CBS | PSP-RS | -3.47E-05 | -1.03659 | 31 | SCP_L Superior_cerebellar_peduncle_left | dti_md_mean | 0.007179 | 0.050255 |
| CBS | PSP-RS | -0.06512 | -1.02534 | 61 | Midbrain_L MIDBRAIN_left | noddi_isovf_mean | 7.34E-08 | 1.47E-07 |
| CBS | PSP-RS | -4.74E-05 | -0.98733 | 57 | GlobusPalidus_L GLOBUS_PALLIDUS_left | dti_md_mean | 0.011042 | 0.088339 |
| CBS | PSP-RS | -0.0471 | -0.95182 | 6 | PrCWM_L PRECENTRAL__WM_left | noddi_icvf_tissue_mean | 1.64E-06 | 1.64E-06 |
| CBS | PSP-RS | -3.56E-05 | -0.95084 | 58 | GlobusPalidus_R GLOBUS_PALLIDUS_right | fwe_md_mean | 8.50E-07 | 1.69E-06 |
| CBS | PSP-RS | 2.61E-05 | 0.950756 | 6 | PrCWM_L PRECENTRAL__WM_left | fwe_md_mean | 8.50E-07 | 1.70E-06 |
| CBS | PSP-RS | 0.401492 | 0.939941 | 57 | GlobusPalidus_L GLOBUS_PALLIDUS_left | fba_fdcount_mean | 1.85E-06 | 7.41E-06 |
| CBS | PSP-RS | -0.03856 | -0.91577 | 7 | PoCWM_L POSTCENTRAL__WM_left | noddi_icvf_tissue_mean | 2.57E-06 | 5.14E-06 |
| CBS | PSP-RS | 2.23E-05 | 0.89416 | 7 | PoCWM_L POSTCENTRAL__WM_left | fwe_md_mean | 3.35E-06 | 1.01E-05 |
| CBS | PSP-RS | 0.494191 | 0.886683 | 55 | Putamen_R PUTAMEN_right | fba_fdcount_mean | 6.99E-06 | 3.49E-05 |
| CBS | PSP-RS | -0.04166 | -0.84296 | 23 | SMWM_L SUPRAMARGINAL__WM_left | noddi_icvf_tissue_mean | 1.57E-05 | 4.72E-05 |
| CBS | PSP-RS | 2.45E-05 | 0.816715 | 23 | SMWM_L SUPRAMARGINAL__WM_left | fwe_md_mean | 2.54E-05 | 0.000102 |
| CBS | PSP-RS | -0.03505 | -0.81606 | 6 | PrCWM_L PRECENTRAL__WM_left | noddi_icvf_mean | 2.07E-05 | 0.000104 |
| CBS | PSP-RS | 0.047552 | 0.79409 | 58 | GlobusPalidus_R GLOBUS_PALLIDUS_right | noddi_icvf_mean | 3.24E-05 | 0.000194 |
| CBS | PSP-RS | -0.02961 | -0.78832 | 69 | PoCWM_R POSTCENTRAL_WM_right | noddi_icvf_tissue_mean | 5.72E-05 | 0.000229 |
| CBS | PSP-RS | 0.030823 | 0.767363 | 108 | PCT_R Pontine_crossing_tract_(a_part_of_MCP)_right | fwe_fa_mean | 0.000307 | 0.000411 |
| CBS | PSP-RS | 0.026492 | 0.762891 | 116 | Midbrain_R MIDBRAIN_right | noddi_icvf_tissue_mean | 7.07E-05 | 0.000465 |
| CBS | PSP-RS | 0.023393 | 0.76088 | 91 | SCP_R Superior_cerebellar_peduncle_right | noddi_icvf_tissue_mean | 7.07E-05 | 0.000492 |
| CBS | PSP-RS | -0.01883 | -0.76072 | 33 | ALIC_L Anterior_limb_of_internal_capsule_left | noddi_isovf_mean | 0.000165 | 0.000494 |
| CBS | PSP-RS | -0.02613 | -0.76064 | 53 | SCC_L Splenium_of_corpus_callosum_left | noddi_icvf_tissue_mean | 7.07E-05 | 0.000495 |
| CBS | PSP-RS | 0.377675 | 0.757221 | 27 | Putamen_L PUTAMEN_left | fba_fdcount_mean | 0.000195 | 0.001167 |
| CBS | PSP-RS | 1.65E-05 | 0.755827 | 69 | PoCWM_R POSTCENTRAL_WM_right | fwe_md_mean | 0.000113 | 0.000565 |
| CBS | PSP-RS | -0.02856 | -0.74998 | 7 | PoCWM_L POSTCENTRAL__WM_left | noddi_icvf_mean | 9.47E-05 | 0.000663 |
| CBS | PSP-RS | -0.02481 | -0.74381 | 62 | Pons_L PONS_left | noddi_odi_mean | 0.000418 | 0.000783 |
| CBS | PSP-RS | -0.02453 | -0.74272 | 69 | PoCWM_R POSTCENTRAL_WM_right | fwe_fa_mean | 0.000307 | 0.000807 |
| CBS | PSP-RS | -0.01016 | -0.74218 | 34 | PLIC_L Posterior_limb_of_internal_capsule_left | noddi_isovf_mean | 0.000205 | 0.000819 |
| CBS | PSP-RS | 0.015413 | 0.741644 | 60 | Thalamus_R THALAMUS_right | noddi_icvf_mean | 0.000104 | 0.000831 |
| CBS | PSP-RS | -0.01755 | -0.73775 | 53 | SCC_L Splenium_of_corpus_callosum_left | fwe_fa_mean | 0.000307 | 0.000922 |
| CBS | PSP-RS | 0.033946 | 0.734355 | 112 | BCC_R Body_of_corpus_callosum_right | noddi_odi_mean | 0.000418 | 0.001011 |
| CBS | PSP-RS | 0.017788 | 0.724459 | 90 | ML_R Medial_lemniscus_right | noddi_icvf_mean | 0.000146 | 0.001316 |
| CBS | PSP-RS | -0.0142 | -0.71631 | 42 | SLF_L Superior_longitudinal_fasciculus_left | noddi_odi_mean | 0.000418 | 0.001633 |
| CBS | PSP-RS | -0.02618 | -0.71538 | 108 | PCT_R Pontine_crossing_tract_(a_part_of_MCP)_right | noddi_odi_mean | 0.000418 | 0.001673 |
| CBS | PSP-RS | 0.312027 | 0.711564 | 61 | Midbrain_L MIDBRAIN_left | fba_fdcount_mean | 0.000523 | 0.003662 |
| CBS | PSP-RS | -0.03175 | -0.69905 | 31 | SCP_L Superior_cerebellar_peduncle_left | noddi_isovf_mean | 0.000423 | 0.002562 |
| CBS | PSP-RS | 0.019462 | 0.69902 | 31 | SCP_L Superior_cerebellar_peduncle_left | fwe_fa_mean | 0.000641 | 0.002564 |
| CBS | PSP-RS | -0.03198 | -0.69655 | 68 | PrCWM_R PRECENTRAL_WM_right | noddi_icvf_tissue_mean | 0.000342 | 0.002733 |

**Table S5** CBS vs PSP-RS

| group_a | group_b | adjusted_mean_diff | cohen_d | roi_id | roi_name | metric | qval_bh | pval_fwe |
| --- | --- | --- | --- | --- | --- | --- | --- | --- |
| PSP-RS | Control | 0.000118 | 3.262822 | 116 | Midbrain_R MIDBRAIN_right | dti_md_mean | 6.77E-19 | 6.77E-19 |
| PSP-RS | Control | 8.08E-05 | 3.10362 | 91 | SCP_R Superior_cerebellar_peduncle_right | dti_md_mean | 4.32E-18 | 8.63E-18 |
| PSP-RS | Control | 7.19E-05 | 2.97994 | 31 | SCP_L Superior_cerebellar_peduncle_left | dti_md_mean | 2.15E-17 | 6.46E-17 |
| PSP-RS | Control | 0.137702 | 2.82255 | 116 | Midbrain_R MIDBRAIN_right | noddi_isovf_mean | 4.59E-49 | 4.59E-49 |
| PSP-RS | Control | 0.000109 | 2.82192 | 61 | Midbrain_L MIDBRAIN_left | dti_md_mean | 2.20E-16 | 8.81E-16 |
| PSP-RS | Control | -0.06201 | -2.63046 | 116 | Midbrain_R MIDBRAIN_right | noddi_icvf_mean | 8.85E-45 | 8.85E-45 |
| PSP-RS | Control | -0.05707 | -2.5576 | 94 | PLIC_R Posterior_limb_of_internal_capsule_right | noddi_icvf_mean | 1.94E-43 | 3.89E-43 |
| PSP-RS | Control | -0.06394 | -2.55225 | 94 | PLIC_R Posterior_limb_of_internal_capsule_right | noddi_icvf_tissue_mean | 5.14E-43 | 5.14E-43 |
| PSP-RS | Control | -0.0604 | -2.5094 | 34 | PLIC_L Posterior_limb_of_internal_capsule_left | noddi_icvf_mean | 1.60E-42 | 4.80E-42 |
| PSP-RS | Control | -0.06488 | -2.38311 | 34 | PLIC_L Posterior_limb_of_internal_capsule_left | noddi_icvf_tissue_mean | 1.80E-39 | 3.60E-39 |
| PSP-RS | Control | 0.126853 | 2.337234 | 61 | Midbrain_L MIDBRAIN_left | noddi_isovf_mean | 2.01E-38 | 4.02E-38 |
| PSP-RS | Control | -0.06211 | -2.29334 | 61 | Midbrain_L MIDBRAIN_left | noddi_icvf_mean | 1.02E-37 | 4.07E-37 |
| PSP-RS | Control | 4.01E-05 | 2.262672 | 90 | ML_R Medial_lemniscus_right | dti_md_mean | 2.49E-12 | 1.24E-11 |
| PSP-RS | Control | 3.62E-05 | 2.227065 | 94 | PLIC_R Posterior_limb_of_internal_capsule_right | fwe_md_mean | 1.35E-35 | 1.35E-35 |
| PSP-RS | Control | 0.081907 | 2.223424 | 31 | SCP_L Superior_cerebellar_peduncle_left | noddi_isovf_mean | 5.45E-36 | 1.63E-35 |
| PSP-RS | Control | -0.05208 | -2.1852 | 3 | SFWM_L SUPERIOR_FRONTAL_WM_left | fwe_fa_mean | 1.23E-34 | 1.23E-34 |
| PSP-RS | Control | 4.02E-05 | 2.180392 | 94 | PLIC_R Posterior_limb_of_internal_capsule_right | dti_md_mean | 8.69E-12 | 5.22E-11 |
| PSP-RS | Control | 3.40E-05 | 2.150978 | 30 | ML_L Medial_lemniscus_left | dti_md_mean | 1.25E-11 | 8.72E-11 |
| PSP-RS | Control | 5.28E-05 | 2.140373 | 117 | Pons_R PONS_right | dti_md_mean | 1.31E-11 | 1.05E-10 |
| PSP-RS | Control | 4.35E-05 | 2.112441 | 34 | PLIC_L Posterior_limb_of_internal_capsule_left | dti_md_mean | 1.90E-11 | 1.71E-10 |
| PSP-RS | Control | -0.04848 | -2.10413 | 3 | SFWM_L SUPERIOR_FRONTAL_WM_left | dti_fa_mean | 1.98E-10 | 1.98E-10 |
| PSP-RS | Control | -0.07802 | -2.08503 | 37 | SCR_L Superior_corona_radiata_left | noddi_icvf_mean | 4.93E-33 | 2.46E-32 |
| PSP-RS | Control | 3.70E-05 | 2.071811 | 34 | PLIC_L Posterior_limb_of_internal_capsule_left | fwe_md_mean | 2.48E-32 | 4.96E-32 |
| PSP-RS | Control | 0.068181 | 2.036646 | 92 | CP_R Cerebral_peduncle_right | noddi_isovf_mean | 7.95E-32 | 3.18E-31 |
| PSP-RS | Control | -0.08192 | -2.02843 | 37 | SCR_L Superior_corona_radiata_left | noddi_icvf_tissue_mean | 1.64E-31 | 4.91E-31 |
| PSP-RS | Control | -0.05914 | -2.02127 | 91 | SCP_R Superior_cerebellar_peduncle_right | dti_fa_mean | 4.20E-10 | 8.40E-10 |
| PSP-RS | Control | 8.36E-05 | 2.003254 | 58 | GlobusPalidus_R GLOBUS_PALLIDUS_right | dti_md_mean | 1.15E-10 | 1.15E-09 |
| PSP-RS | Control | -0.07428 | -1.99912 | 97 | SCR_R Superior_corona_radiata_right | noddi_icvf_mean | 3.84E-31 | 2.31E-30 |
| PSP-RS | Control | -0.06232 | -1.98105 | 31 | SCP_L Superior_cerebellar_peduncle_left | dti_fa_mean | 5.65E-10 | 1.70E-09 |
| PSP-RS | Control | -0.06245 | -1.97869 | 6 | PrCWM_L PRECENTRAL__WM_left | noddi_icvf_mean | 9.68E-31 | 6.77E-30 |
| PSP-RS | Control | 5.30E-05 | 1.976858 | 62 | Pons_L PONS_left | dti_md_mean | 1.66E-10 | 1.82E-09 |
| PSP-RS | Control | -0.07761 | -1.96173 | 4 | MFWM_L MIDDLE_FRONTAL__WM_left | noddi_icvf_tissue_mean | 4.14E-30 | 1.66E-29 |
| PSP-RS | Control | -0.06068 | -1.95314 | 68 | PrCWM_R PRECENTRAL_WM_right | noddi_icvf_mean | 3.26E-30 | 2.60E-29 |
| PSP-RS | Control | -0.07865 | -1.95314 | 97 | SCR_R Superior_corona_radiata_right | noddi_icvf_tissue_mean | 5.21E-30 | 2.60E-29 |
| PSP-RS | Control | -0.07263 | -1.88905 | 4 | MFWM_L MIDDLE_FRONTAL__WM_left | noddi_icvf_mean | 8.42E-29 | 7.58E-28 |
| PSP-RS | Control | -0.05236 | -1.88138 | 91 | SCP_R Superior_cerebellar_peduncle_right | noddi_icvf_mean | 1.13E-28 | 1.13E-27 |
| PSP-RS | Control | -0.05946 | -1.87633 | 65 | SFWM_R SUPERIOR_FRONTAL__WM_right | noddi_icvf_mean | 1.34E-28 | 1.48E-27 |
| PSP-RS | Control | -0.0501 | -1.87173 | 31 | SCP_L Superior_cerebellar_peduncle_left | noddi_icvf_mean | 1.57E-28 | 1.88E-27 |
| PSP-RS | Control | 0.085713 | 1.841677 | 91 | SCP_R Superior_cerebellar_peduncle_right | noddi_isovf_mean | 1.82E-27 | 9.08E-27 |
| PSP-RS | Control | -0.06399 | -1.83619 | 68 | PrCWM_R PRECENTRAL_WM_right | noddi_icvf_tissue_mean | 2.02E-27 | 1.21E-26 |
| PSP-RS | Control | -0.04879 | -1.81954 | 65 | SFWM_R SUPERIOR_FRONTAL__WM_right | fwe_fa_mean | 1.44E-26 | 2.89E-26 |
| PSP-RS | Control | -0.06516 | -1.81321 | 6 | PrCWM_L PRECENTRAL__WM_left | noddi_icvf_tissue_mean | 5.74E-27 | 4.02E-26 |
| PSP-RS | Control | 4.32E-05 | 1.810306 | 29 | ICP_L Inferior_cerebellar_peduncle_left | dti_md_mean | 2.76E-09 | 3.31E-08 |
| PSP-RS | Control | 3.29E-05 | 1.791017 | 68 | PrCWM_R PRECENTRAL_WM_right | fwe_md_mean | 4.26E-26 | 1.28E-25 |
| PSP-RS | Control | 0.061528 | 1.788131 | 32 | CP_L Cerebral_peduncle_left | noddi_isovf_mean | 2.48E-26 | 1.49E-25 |
| PSP-RS | Control | -0.06297 | -1.78796 | 65 | SFWM_R SUPERIOR_FRONTAL__WM_right | noddi_icvf_tissue_mean | 1.87E-26 | 1.50E-25 |
| PSP-RS | Control | -0.04545 | -1.78295 | 65 | SFWM_R SUPERIOR_FRONTAL__WM_right | dti_fa_mean | 1.33E-08 | 5.32E-08 |
| PSP-RS | Control | 4.24E-05 | 1.774429 | 4 | MFWM_L MIDDLE_FRONTAL__WM_left | fwe_md_mean | 6.53E-26 | 3.03E-25 |
| PSP-RS | Control | 3.34E-05 | 1.773004 | 6 | PrCWM_L PRECENTRAL__WM_left | fwe_md_mean | 6.53E-26 | 3.27E-25 |
| PSP-RS | Control | -0.05763 | -1.76945 | 3 | SFWM_L SUPERIOR_FRONTAL_WM_left | noddi_icvf_mean | 3.02E-26 | 3.93E-25 |

**Table S6** PSP-RS vs Controls

| group_a | group_b | adjusted_mean_diff | cohen_d | roi_id | roi_name | metric | qval_bh | pval_fwe |
| --- | --- | --- | --- | --- | --- | --- | --- | --- |
| PSP-RS | PD | 6.07E-05 | 2.244094 | 31 | SCP_L Superior_cerebellar_peduncle_left | dti_md_mean | 2.32E-07 | 2.32E-07 |
| PSP-RS | PD | 6.48E-05 | 2.081059 | 91 | SCP_R Superior_cerebellar_peduncle_right | dti_md_mean | 8.49E-07 | 1.70E-06 |
| PSP-RS | PD | 8.92E-05 | 1.936261 | 61 | Midbrain_L MIDBRAIN_left | dti_md_mean | 3.28E-06 | 9.84E-06 |
| PSP-RS | PD | 8.88E-05 | 1.908695 | 116 | Midbrain_R MIDBRAIN_right | dti_md_mean | 3.43E-06 | 1.37E-05 |
| PSP-RS | PD | 0.071707 | 1.759647 | 31 | SCP_L Superior_cerebellar_peduncle_left | noddi_isovf_mean | 5.21E-16 | 5.21E-16 |
| PSP-RS | PD | -0.04626 | -1.75121 | 116 | Midbrain_R MIDBRAIN_right | noddi_icvf_mean | 7.03E-16 | 7.03E-16 |
| PSP-RS | PD | -0.03771 | -1.70226 | 3 | SFWM_L SUPERIOR_FRONTAL_WM_left | dti_fa_mean | 0.00016 | 0.00016 |
| PSP-RS | PD | -0.04996 | -1.65441 | 61 | Midbrain_L MIDBRAIN_left | noddi_icvf_mean | 1.05E-14 | 2.10E-14 |
| PSP-RS | PD | -0.03839 | -1.62427 | 3 | SFWM_L SUPERIOR_FRONTAL_WM_left | fwe_fa_mean | 5.98E-14 | 5.98E-14 |
| PSP-RS | PD | 0.101981 | 1.58487 | 116 | Midbrain_R MIDBRAIN_right | noddi_isovf_mean | 1.16E-13 | 2.32E-13 |
| PSP-RS | PD | 0.104407 | 1.566678 | 61 | Midbrain_L MIDBRAIN_left | noddi_isovf_mean | 1.44E-13 | 4.31E-13 |
| PSP-RS | PD | -0.0383 | -1.51914 | 65 | SFWM_R SUPERIOR_FRONTAL__WM_right | dti_fa_mean | 0.000659 | 0.001319 |
| PSP-RS | PD | -0.04012 | -1.48964 | 65 | SFWM_R SUPERIOR_FRONTAL__WM_right | fwe_fa_mean | 2.89E-12 | 5.78E-12 |
| PSP-RS | PD | -0.04608 | -1.46106 | 91 | SCP_R Superior_cerebellar_peduncle_right | noddi_icvf_mean | 4.98E-12 | 1.49E-11 |
| PSP-RS | PD | -0.05065 | -1.42199 | 31 | SCP_L Superior_cerebellar_peduncle_left | dti_fa_mean | 0.00101 | 0.003903 |
| PSP-RS | PD | -0.05031 | -1.41881 | 91 | SCP_R Superior_cerebellar_peduncle_right | dti_fa_mean | 0.00101 | 0.004042 |
| PSP-RS | PD | 2.88E-05 | 1.405621 | 90 | ML_R Medial_lemniscus_right | dti_md_mean | 0.000935 | 0.004673 |
| PSP-RS | PD | -0.04035 | -1.3943 | 94 | PLIC_R Posterior_limb_of_internal_capsule_right | noddi_icvf_tissue_mean | 1.32E-10 | 1.32E-10 |
| PSP-RS | PD | 3.86E-05 | 1.301366 | 117 | Pons_R PONS_right | dti_md_mean | 0.002309 | 0.014404 |
| PSP-RS | PD | -0.03221 | -1.29342 | 6 | PrCWM_L PRECENTRAL__WM_left | dti_fa_mean | 0.003134 | 0.015669 |
| PSP-RS | PD | 6.27E-05 | 1.290493 | 58 | GlobusPalidus_R GLOBUS_PALLIDUS_right | dti_md_mean | 0.002309 | 0.016162 |
| PSP-RS | PD | 0.049849 | 1.281634 | 32 | CP_L Cerebral_peduncle_left | noddi_isovf_mean | 1.17E-09 | 4.69E-09 |
| PSP-RS | PD | -0.0387 | -1.25981 | 34 | PLIC_L Posterior_limb_of_internal_capsule_left | noddi_icvf_tissue_mean | 4.60E-09 | 9.20E-09 |
| PSP-RS | PD | -0.03444 | -1.25645 | 94 | PLIC_R Posterior_limb_of_internal_capsule_right | noddi_icvf_mean | 2.55E-09 | 1.02E-08 |
| PSP-RS | PD | -0.03632 | -1.22135 | 31 | SCP_L Superior_cerebellar_peduncle_left | noddi_icvf_mean | 5.92E-09 | 2.96E-08 |
| PSP-RS | PD | -0.40557 | -1.20591 | 58 | GlobusPalidus_R GLOBUS_PALLIDUS_right | fba_fdcount_mean | 1.51E-06 | 1.51E-06 |
| PSP-RS | PD | 0.066074 | 1.203581 | 91 | SCP_R Superior_cerebellar_peduncle_right | noddi_isovf_mean | 1.01E-08 | 5.05E-08 |
| PSP-RS | PD | -0.04053 | -1.19658 | 6 | PrCWM_L PRECENTRAL__WM_left | noddi_icvf_mean | 1.04E-08 | 6.22E-08 |
| PSP-RS | PD | 0.048656 | 1.195026 | 92 | CP_R Cerebral_peduncle_right | noddi_isovf_mean | 1.09E-08 | 6.52E-08 |
| PSP-RS | PD | 3.24E-05 | 1.194648 | 6 | PrCWM_L PRECENTRAL__WM_left | dti_md_mean | 0.005465 | 0.043723 |
| PSP-RS | PD | -0.03858 | -1.18272 | 65 | SFWM_R SUPERIOR_FRONTAL__WM_right | noddi_icvf_mean | 1.34E-08 | 9.39E-08 |
| PSP-RS | PD | -0.03992 | -1.17108 | 68 | PrCWM_R PRECENTRAL_WM_right | noddi_icvf_mean | 1.66E-08 | 1.32E-07 |
| PSP-RS | PD | -0.0317 | -1.14867 | 6 | PrCWM_L PRECENTRAL__WM_left | fwe_fa_mean | 8.50E-08 | 2.55E-07 |
| PSP-RS | PD | -0.03462 | -1.1421 | 34 | PLIC_L Posterior_limb_of_internal_capsule_left | noddi_icvf_mean | 3.43E-08 | 3.09E-07 |
| PSP-RS | PD | 2.20E-05 | 1.138993 | 94 | PLIC_R Posterior_limb_of_internal_capsule_right | fwe_md_mean | 3.38E-07 | 3.38E-07 |
| PSP-RS | PD | 2.96E-05 | 1.12453 | 68 | PrCWM_R PRECENTRAL_WM_right | dti_md_mean | 0.009818 | 0.088358 |
| PSP-RS | PD | -0.04256 | -1.10926 | 66 | MFWM_R MIDDLE_FRONTAL_WM_right | noddi_icvf_mean | 7.93E-08 | 7.93E-07 |
| PSP-RS | PD | -0.03317 | -1.10365 | 37 | SCR_L Superior_corona_radiata_left | dti_fa_mean | 0.01808 | 0.108482 |
| PSP-RS | PD | 3.00E-05 | 1.102949 | 65 | SFWM_R SUPERIOR_FRONTAL__WM_right | dti_md_mean | 0.010923 | 0.109229 |
| PSP-RS | PD | -0.04431 | -1.08773 | 108 | PCT_R Pontine_crossing_tract_(a_part_of_MCP)_right | fwe_fa_mean | 3.65E-07 | 1.46E-06 |
| PSP-RS | PD | 0.041851 | 1.078306 | 41 | Fx_ST_L Fornix(cres)_Stria_terminalisleft | noddi_icvf_tissue_mean | 6.33E-07 | 1.90E-06 |
| PSP-RS | PD | -0.02157 | -1.0726 | 116 | Midbrain_R MIDBRAIN_right | dti_fa_mean | 0.020947 | 0.146626 |
| PSP-RS | PD | 2.21E-05 | 1.068087 | 68 | PrCWM_R PRECENTRAL_WM_right | fwe_md_mean | 1.26E-06 | 2.53E-06 |
| PSP-RS | PD | 3.55E-05 | 1.053483 | 66 | MFWM_R MIDDLE_FRONTAL_WM_right | dti_md_mean | 0.016009 | 0.176099 |
| PSP-RS | PD | -0.03819 | -1.05338 | 65 | SFWM_R SUPERIOR_FRONTAL__WM_right | noddi_icvf_tissue_mean | 9.49E-07 | 3.79E-06 |
| PSP-RS | PD | -0.03552 | -1.05298 | 3 | SFWM_L SUPERIOR_FRONTAL_WM_left | noddi_icvf_mean | 3.49E-07 | 3.84E-06 |
| PSP-RS | PD | -0.04888 | -1.04778 | 52 | BCC_L Body_of_corpus_callosum_left | dti_fa_mean | 0.022059 | 0.185923 |
| PSP-RS | PD | 3.37E-05 | 1.043033 | 62 | Pons_L PONS_left | dti_md_mean | 0.016208 | 0.194495 |
| PSP-RS | PD | -0.03224 | -1.04087 | 1 | SPWM_L SUPERIOR_PARIETAL__WM_left | dti_fa_mean | 0.022059 | 0.198527 |
| PSP-RS | PD | -0.03486 | -1.03833 | 61 | Midbrain_L MIDBRAIN_left | noddi_icvf_tissue_mean | 1.14E-06 | 5.73E-06 |

**Table S7** PSP-RS vs PD

| group_a | group_b | adjusted_mean_diff | cohen_d | roi_id | roi_name | metric | qval_bh | pval_fwe |
| --- | --- | --- | --- | --- | --- | --- | --- | --- |
| PD | Control | 1.99E-05 | 1.386769 | 79 | STWM_R SUPERIOR_TEMPORAL_WM_right | fwe_md_mean | 3.82E-12 | 3.82E-12 |
| PD | Control | -0.03268 | -1.34937 | 45 | SS_L Sagittal_stratum__left | noddi_icvf_mean | 1.44E-11 | 1.44E-11 |
| PD | Control | 2.14E-05 | 1.334227 | 46 | EC_L External_capsule_left | fwe_md_mean | 9.72E-12 | 2.44E-11 |
| PD | Control | 1.63E-05 | 1.329175 | 65 | SFWM_R SUPERIOR_FRONTAL__WM_right | fwe_md_mean | 9.72E-12 | 2.92E-11 |
| PD | Control | -0.03204 | -1.31886 | 105 | SS_R Sagittal_stratum_right | noddi_icvf_mean | 1.60E-11 | 4.18E-11 |
| PD | Control | 3.11E-05 | 1.31627 | 105 | SS_R Sagittal_stratum_right | dti_md_mean | 0.001458 | 0.003712 |
| PD | Control | -0.03301 | -1.31499 | 64 | SPWM_R SUPERIOR_PARIETAL_WM_right | noddi_icvf_mean | 1.60E-11 | 4.79E-11 |
| PD | Control | 1.64E-05 | 1.304263 | 3 | SFWM_L SUPERIOR_FRONTAL_WM_left | fwe_md_mean | 1.74E-11 | 6.95E-11 |
| PD | Control | -0.02536 | -1.3029 | 3 | SFWM_L SUPERIOR_FRONTAL_WM_left | noddi_icvf_mean | 1.82E-11 | 7.29E-11 |
| PD | Control | 2.95E-05 | 1.294208 | 105 | SS_R Sagittal_stratum_right | fwe_md_mean | 1.97E-11 | 9.85E-11 |
| PD | Control | 2.21E-05 | 1.292213 | 79 | STWM_R SUPERIOR_TEMPORAL_WM_right | dti_md_mean | 0.001458 | 0.004923 |
| PD | Control | 1.87E-05 | 1.285863 | 3 | SFWM_L SUPERIOR_FRONTAL_WM_left | dti_md_mean | 0.001458 | 0.005302 |
| PD | Control | -0.02931 | -1.27118 | 22 | MFOWM_L MIDDLE_FRONTO-ORBITAL__WM_left | noddi_icvf_mean | 4.36E-11 | 2.18E-10 |
| PD | Control | 2.97E-05 | 1.265036 | 45 | SS_L Sagittal_stratum__left | dti_md_mean | 0.001458 | 0.006755 |
| PD | Control | -0.02498 | -1.2507 | 65 | SFWM_R SUPERIOR_FRONTAL__WM_right | noddi_icvf_mean | 6.56E-11 | 4.39E-10 |
| PD | Control | -0.03078 | -1.24938 | 1 | SPWM_L SUPERIOR_PARIETAL__WM_left | noddi_icvf_mean | 6.56E-11 | 4.59E-10 |
| PD | Control | -0.03809 | -1.23952 | 42 | SLF_L Superior_longitudinal_fasciculus_left | noddi_icvf_mean | 7.46E-11 | 6.42E-10 |
| PD | Control | -0.04033 | -1.23819 | 102 | SLF_R Superior_longitudinal_fasciculus_right | noddi_icvf_mean | 7.46E-11 | 6.72E-10 |
| PD | Control | -0.04421 | -1.23538 | 42 | SLF_L Superior_longitudinal_fasciculus_left | noddi_icvf_tissue_mean | 7.23E-10 | 7.39E-10 |
| PD | Control | 1.85E-05 | 1.235281 | 65 | SFWM_R SUPERIOR_FRONTAL__WM_right | dti_md_mean | 0.001458 | 0.009525 |
| PD | Control | 2.64E-05 | 1.235272 | 24 | RWM_L RECTUS__WM_left | dti_md_mean | 0.001458 | 0.009526 |
| PD | Control | 2.57E-05 | 1.229268 | 22 | MFOWM_L MIDDLE_FRONTO-ORBITAL__WM_left | dti_md_mean | 0.001458 | 0.010206 |
| PD | Control | -0.02838 | -1.21551 | 3 | SFWM_L SUPERIOR_FRONTAL_WM_left | noddi_icvf_tissue_mean | 7.23E-10 | 1.45E-09 |
| PD | Control | -0.03728 | -1.20645 | 4 | MFWM_L MIDDLE_FRONTAL__WM_left | noddi_icvf_mean | 1.96E-10 | 1.96E-09 |
| PD | Control | 1.95E-05 | 1.196805 | 1 | SPWM_L SUPERIOR_PARIETAL__WM_left | fwe_md_mean | 4.51E-10 | 2.71E-09 |
| PD | Control | 2.45E-05 | 1.188971 | 64 | SPWM_R SUPERIOR_PARIETAL_WM_right | dti_md_mean | 0.001845 | 0.016153 |
| PD | Control | 2.20E-05 | 1.186545 | 1 | SPWM_L SUPERIOR_PARIETAL__WM_left | dti_md_mean | 0.001845 | 0.016602 |
| PD | Control | -0.04557 | -1.18577 | 102 | SLF_R Superior_longitudinal_fasciculus_right | noddi_icvf_tissue_mean | 1.30E-09 | 3.91E-09 |
| PD | Control | 1.85E-05 | 1.183956 | 34 | PLIC_L Posterior_limb_of_internal_capsule_left | fwe_md_mean | 5.21E-10 | 4.15E-09 |
| PD | Control | 2.03E-05 | 1.182348 | 20 | MTWM_L MIDDLE_TEMPORAL__WM_left | fwe_md_mean | 5.21E-10 | 4.38E-09 |
| PD | Control | 2.00E-05 | 1.180287 | 81 | MTWM_R MIDDLE_TEMPORAL_WM_right | fwe_md_mean | 5.21E-10 | 4.69E-09 |
| PD | Control | -0.02695 | -1.1729 | 95 | PTR_R Posterior_thalamic_radiation_right | dti_fa_mean | 0.019365 | 0.019365 |
| PD | Control | 1.80E-05 | 1.172603 | 64 | SPWM_R SUPERIOR_PARIETAL_WM_right | fwe_md_mean | 5.50E-10 | 6.04E-09 |
| PD | Control | 1.91E-05 | 1.172565 | 18 | STWM_L SUPERIOR_TEMPORAL__WM_left | fwe_md_mean | 5.50E-10 | 6.05E-09 |
| PD | Control | 1.57E-05 | 1.164649 | 69 | PoCWM_R POSTCENTRAL_WM_right | fwe_md_mean | 6.55E-10 | 7.86E-09 |
| PD | Control | -0.02665 | -1.1634 | 46 | EC_L External_capsule_left | noddi_icvf_mean | 6.87E-10 | 8.19E-09 |
| PD | Control | -0.02509 | -1.16321 | 79 | STWM_R SUPERIOR_TEMPORAL_WM_right | noddi_icvf_mean | 6.87E-10 | 8.24E-09 |
| PD | Control | -0.03568 | -1.16304 | 1 | SPWM_L SUPERIOR_PARIETAL__WM_left | noddi_icvf_tissue_mean | 1.53E-09 | 8.29E-09 |
| PD | Control | -0.03795 | -1.15774 | 23 | SMWM_L SUPRAMARGINAL__WM_left | noddi_icvf_tissue_mean | 1.53E-09 | 9.86E-09 |
| PD | Control | -0.03514 | -1.15734 | 64 | SPWM_R SUPERIOR_PARIETAL_WM_right | noddi_icvf_tissue_mean | 1.53E-09 | 9.99E-09 |
| PD | Control | -0.02751 | -1.15533 | 65 | SFWM_R SUPERIOR_FRONTAL__WM_right | noddi_icvf_tissue_mean | 1.53E-09 | 1.07E-08 |
| PD | Control | -0.0262 | -1.1522 | 34 | PLIC_L Posterior_limb_of_internal_capsule_left | noddi_icvf_mean | 9.10E-10 | 1.18E-08 |
| PD | Control | 0.039459 | 1.150716 | 88 | CST_R Corticospinal_tract_right | noddi_isovf_mean | 1.24E-08 | 1.24E-08 |
| PD | Control | 2.15E-05 | 1.150146 | 20 | MTWM_L MIDDLE_TEMPORAL__WM_left | dti_md_mean | 0.002368 | 0.024987 |
| PD | Control | 2.22E-05 | 1.148437 | 4 | MFWM_L MIDDLE_FRONTAL__WM_left | fwe_md_mean | 1.03E-09 | 1.34E-08 |
| PD | Control | 2.15E-05 | 1.142965 | 81 | MTWM_R MIDDLE_TEMPORAL_WM_right | dti_md_mean | 0.002368 | 0.027069 |
| PD | Control | -0.03682 | -1.14183 | 35 | PTR_L Posterior_thalamic_radiation_left | noddi_icvf_mean | 1.19E-09 | 1.66E-08 |
| PD | Control | -0.02958 | -1.14123 | 54 | RLIC_L Retrolenticular_part_of_internal_capsule_left | noddi_icvf_tissue_mean | 2.12E-09 | 1.69E-08 |
| PD | Control | 2.73E-05 | 1.133063 | 46 | EC_L External_capsule_left | dti_md_mean | 0.002368 | 0.030215 |
| PD | Control | -0.03399 | -1.1324 | 95 | PTR_R Posterior_thalamic_radiation_right | noddi_icvf_mean | 1.43E-09 | 2.26E-08 |

**Table S8** PD vs Controls

#### Whole cohort and within disease group Correlation

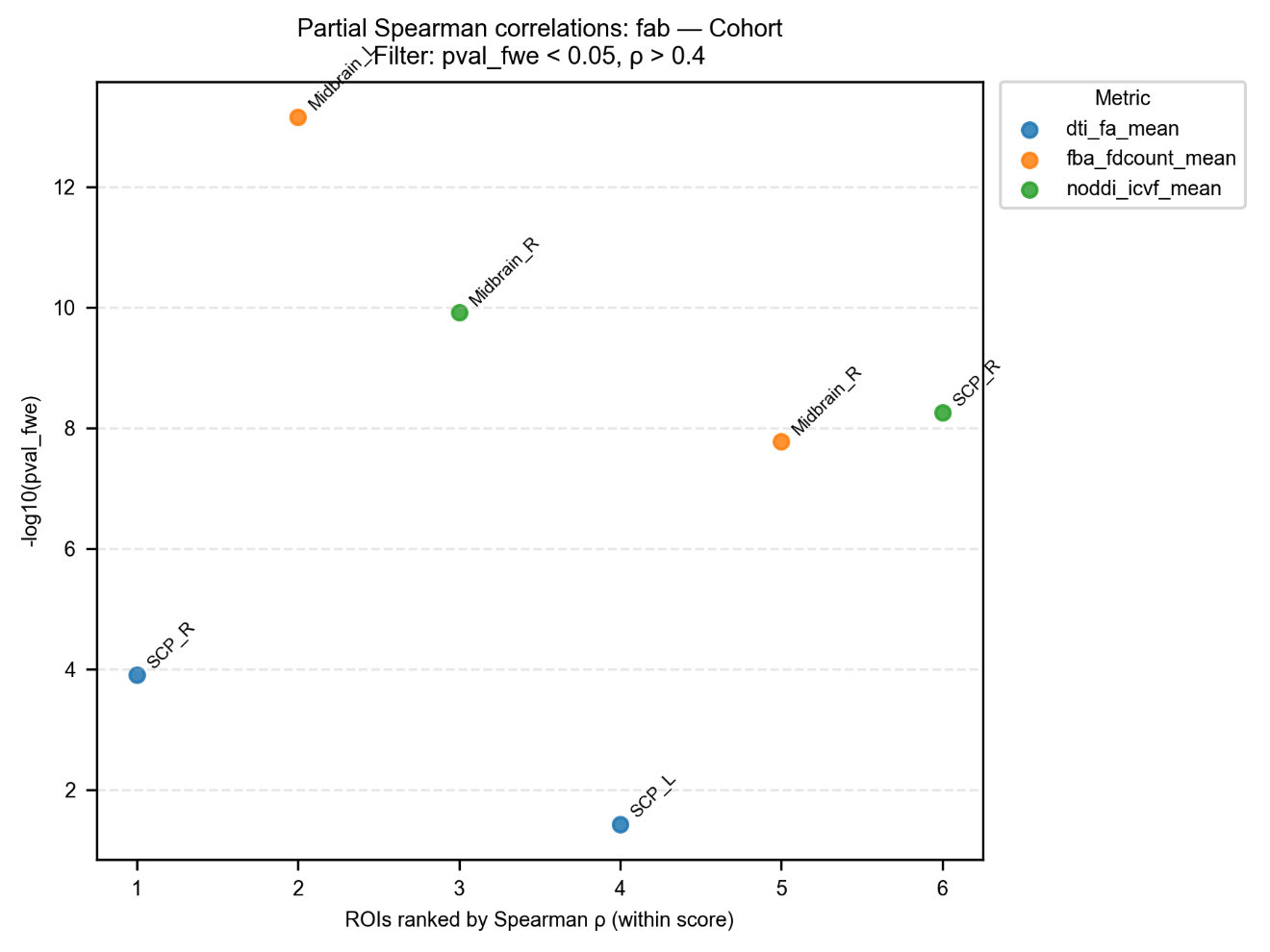

Figure S8 Correlating with FAB in cohort.

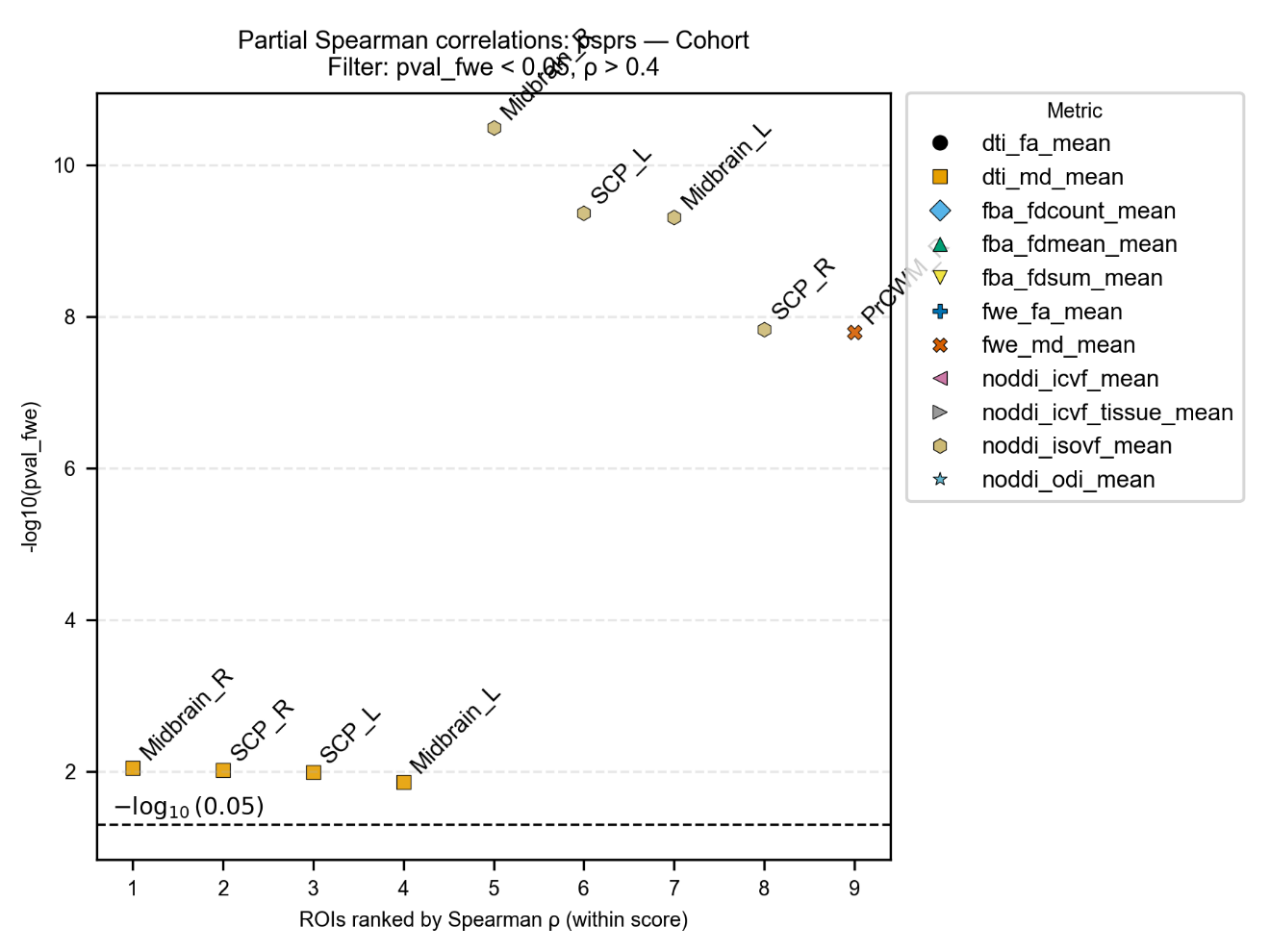

Figure S9 Correlating with PSP Rating Scale in cohort.

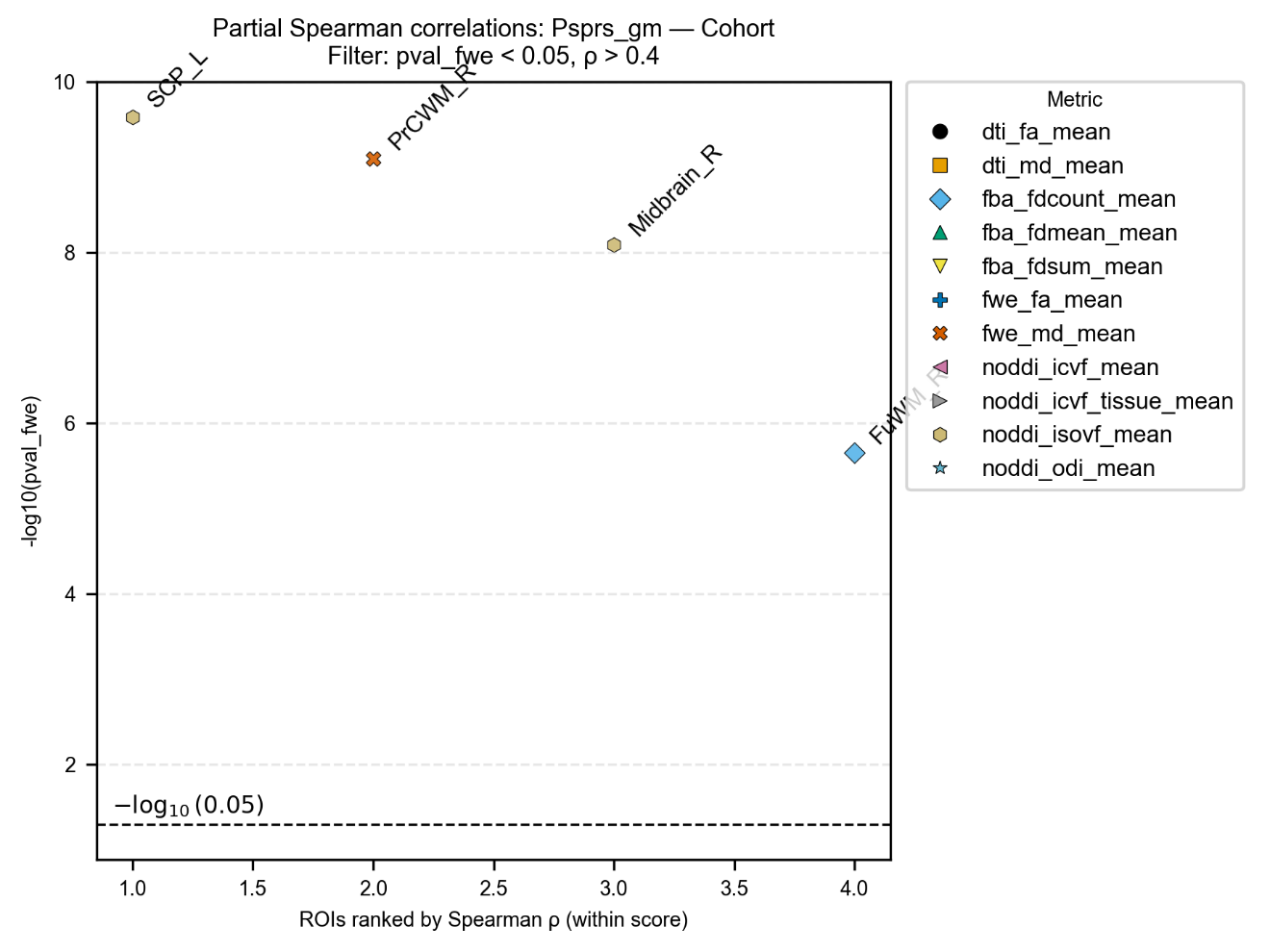

Figure S10 Correlating with PSP Rating Scale Gait/midline in cohort.

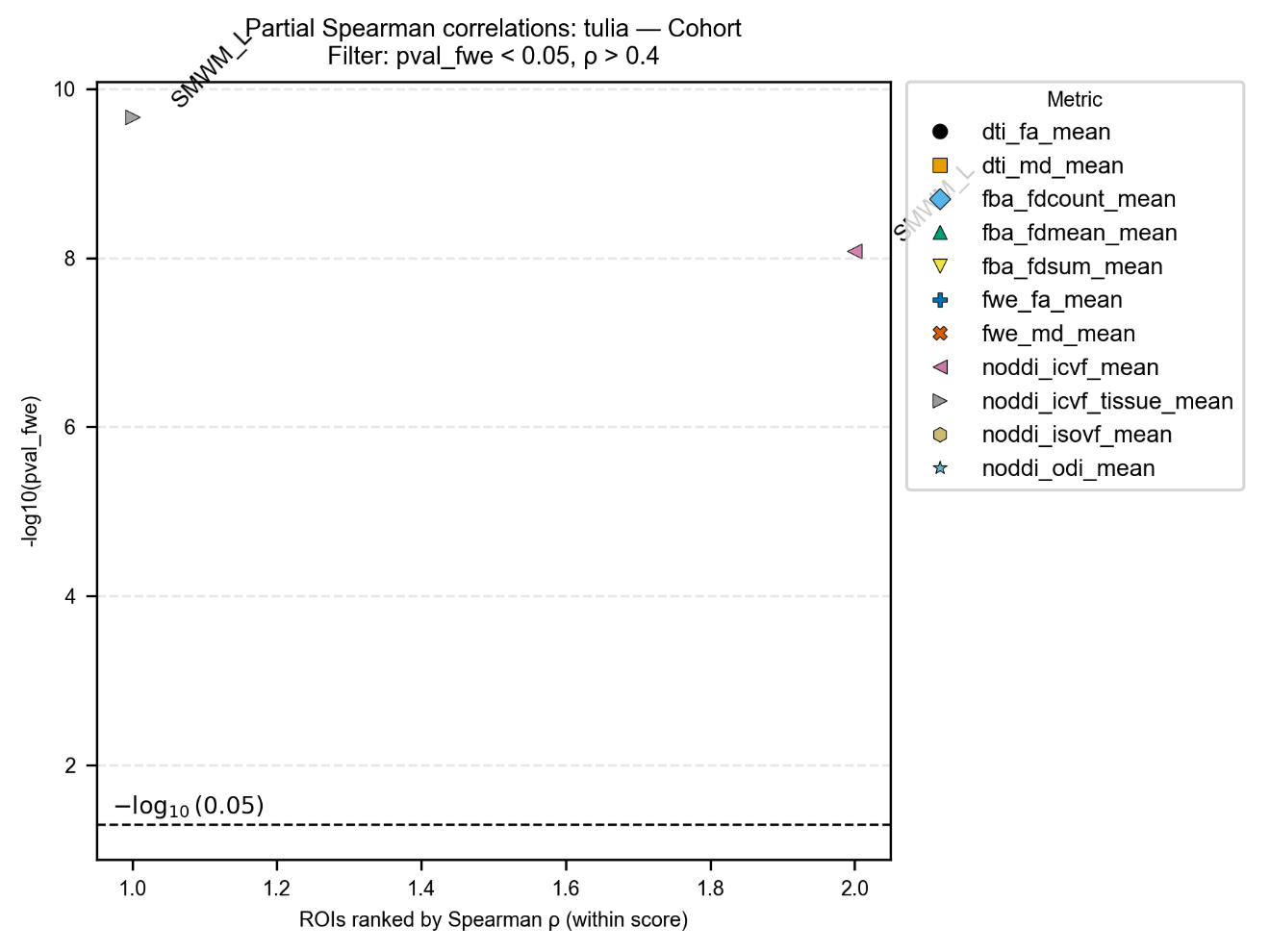

Figure S11 Correlating with TULIA in cohort.

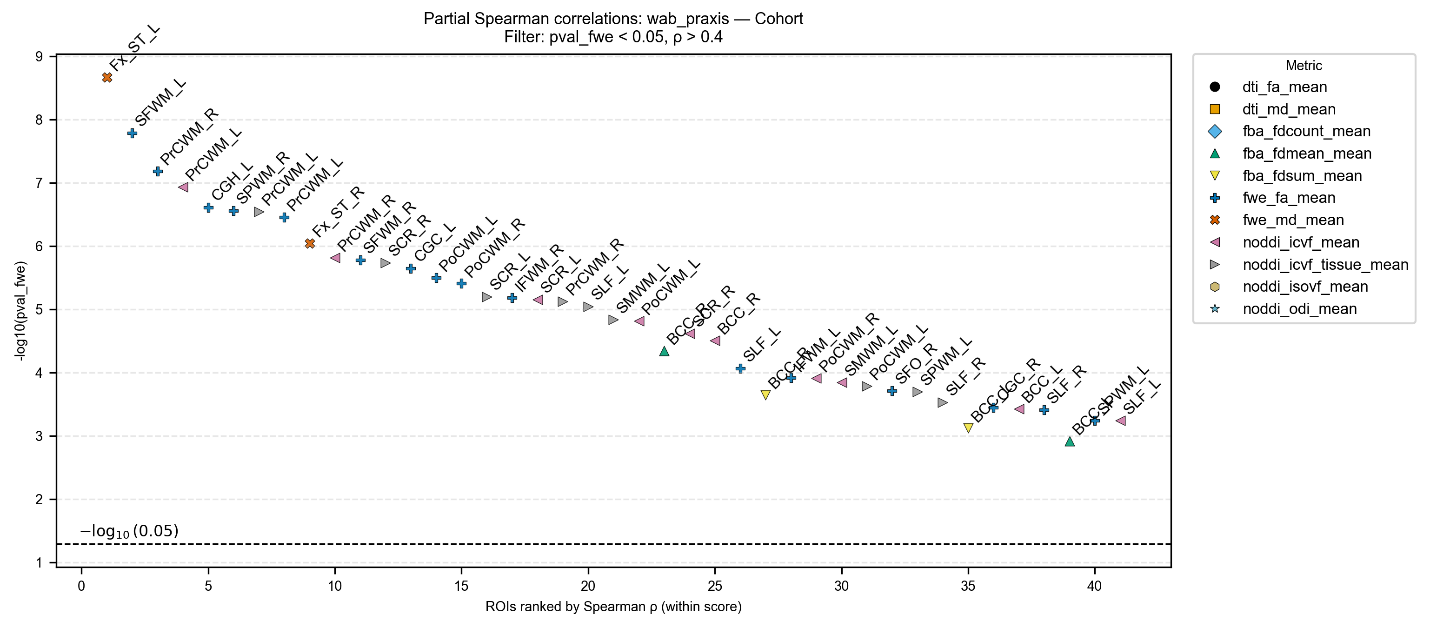

Figure S12 Correlating with WAB-Praxis in cohort.

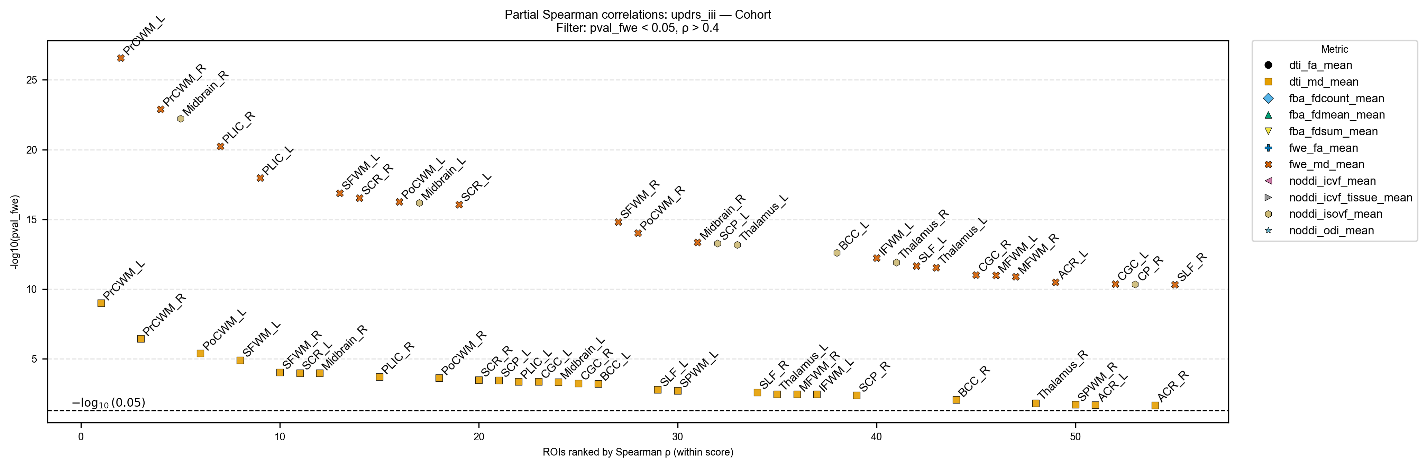

Figure S13 Correlating with MDS-UPDRS III in cohort.

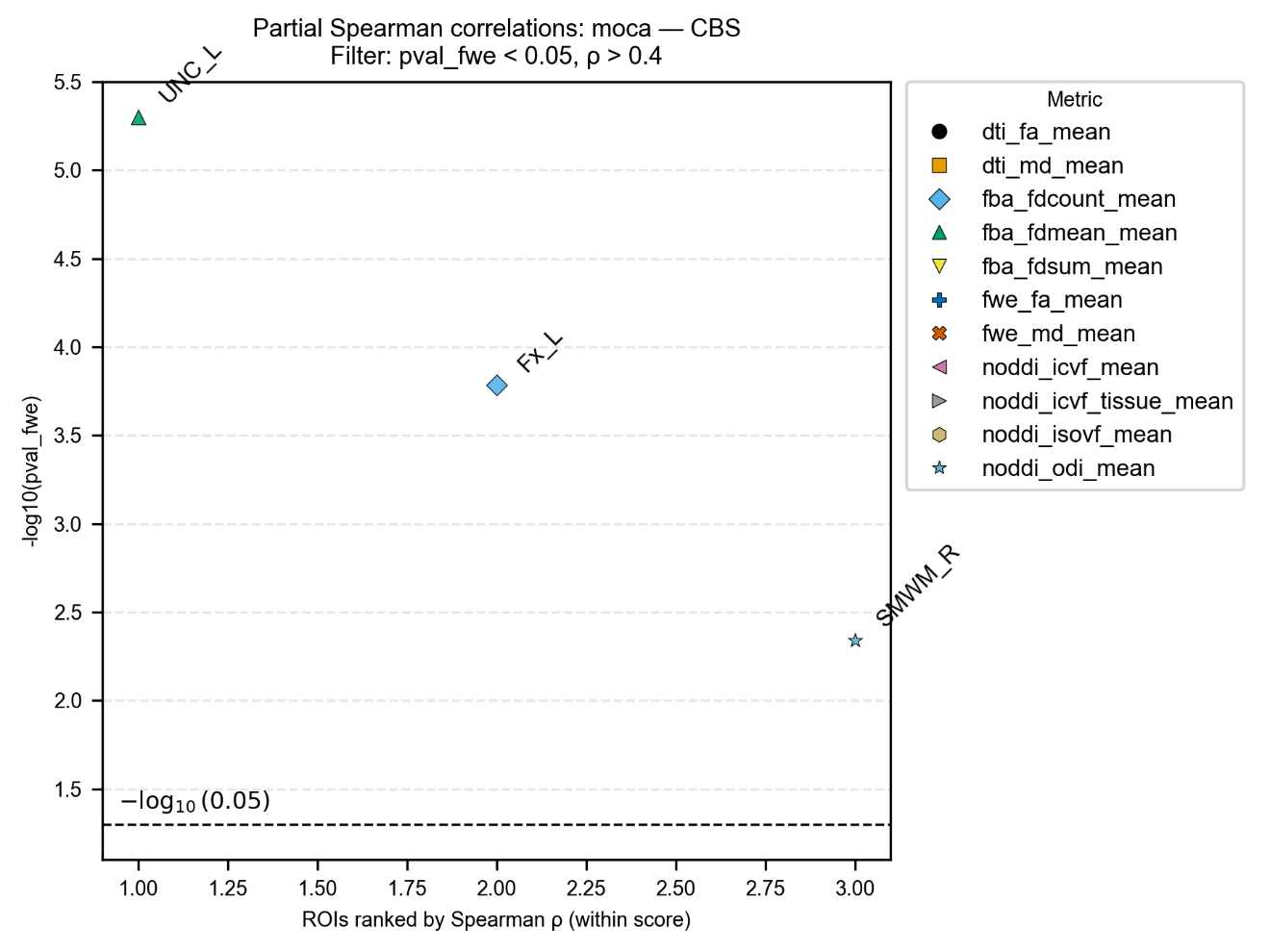

Figure S14 Correlating with MoCA in CBS group.

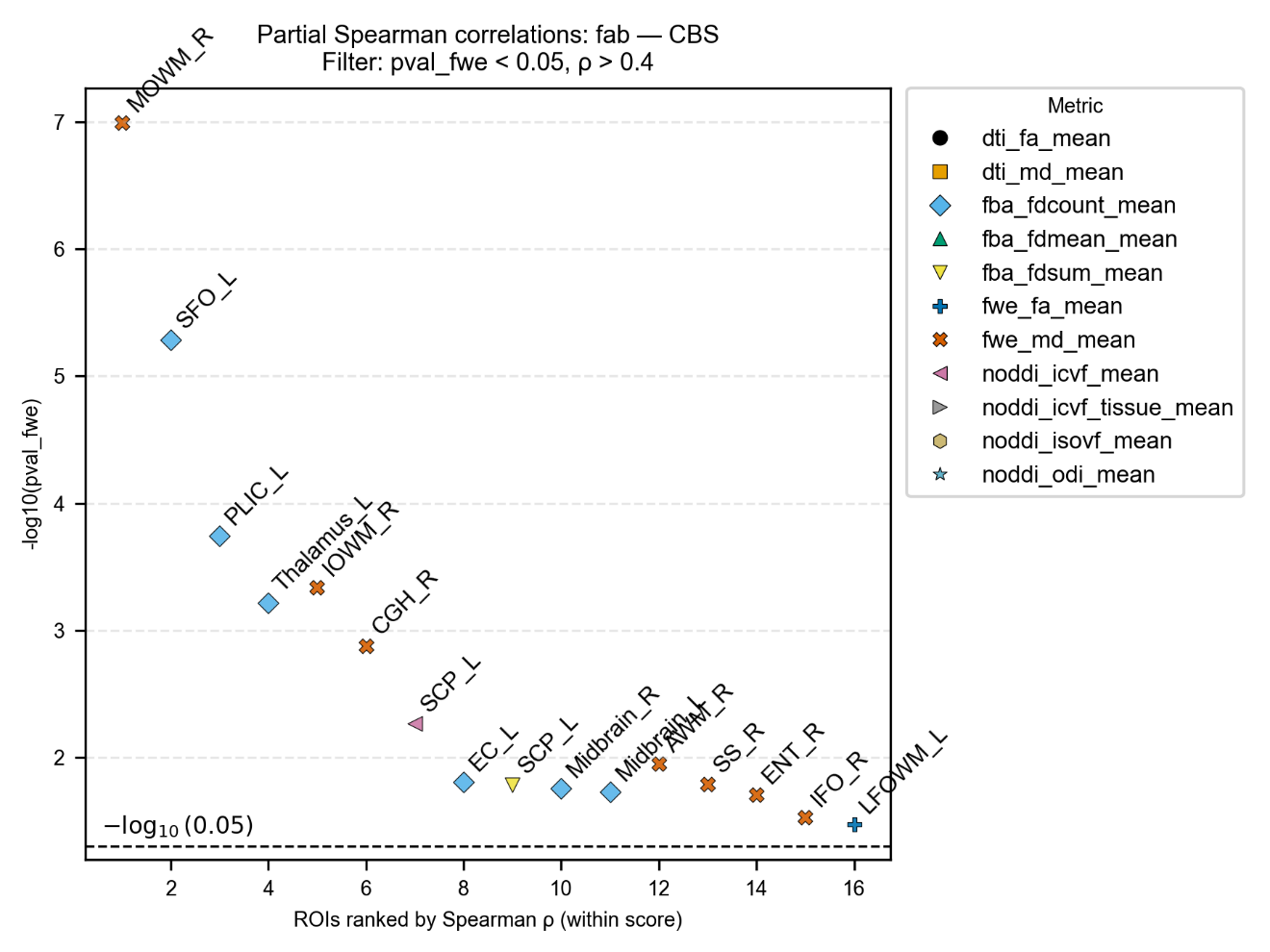

Figure S15 Correlating with FAB in CBS group.

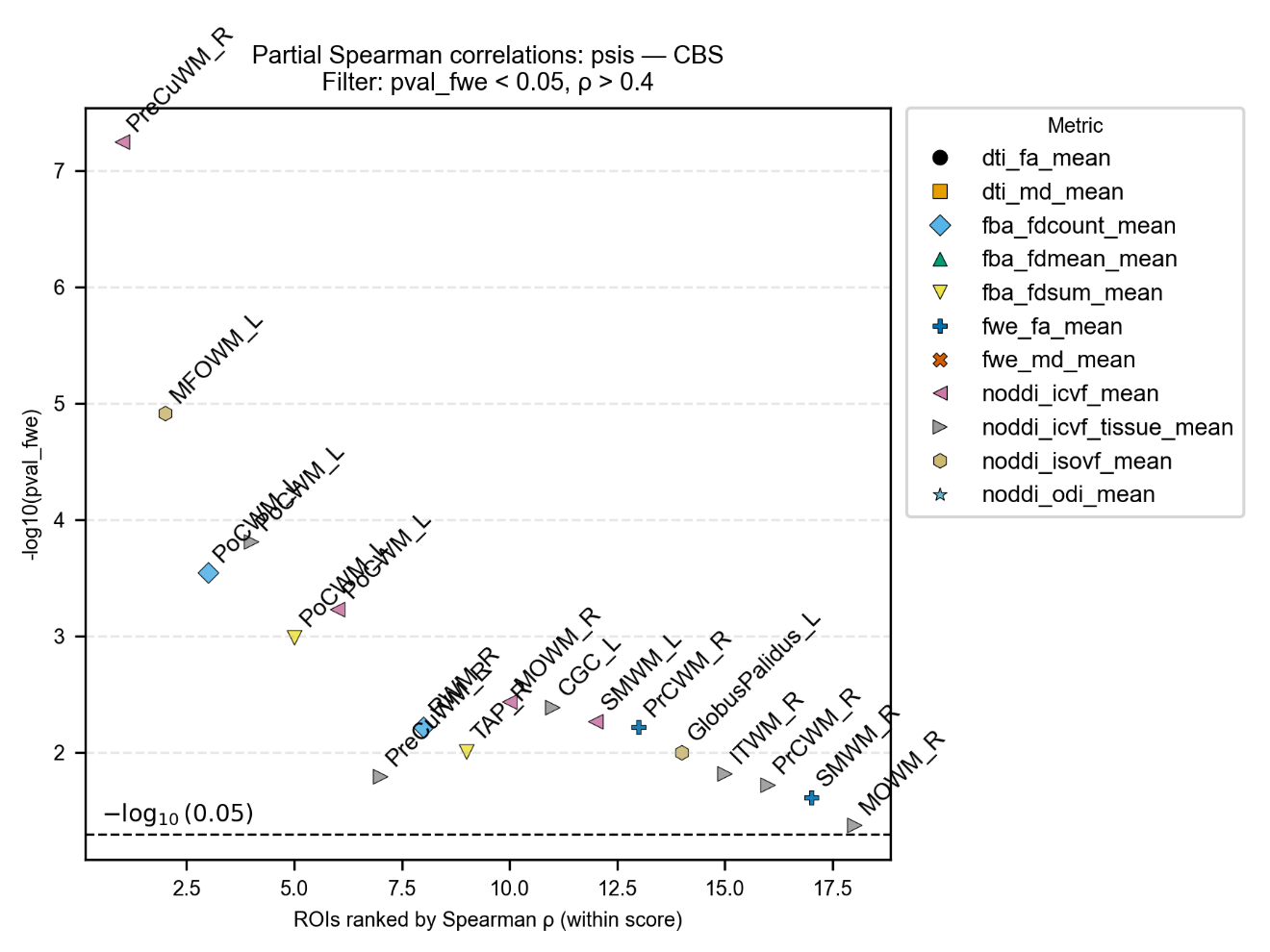

Figure S16 Correlating with PSIS in CBS group.

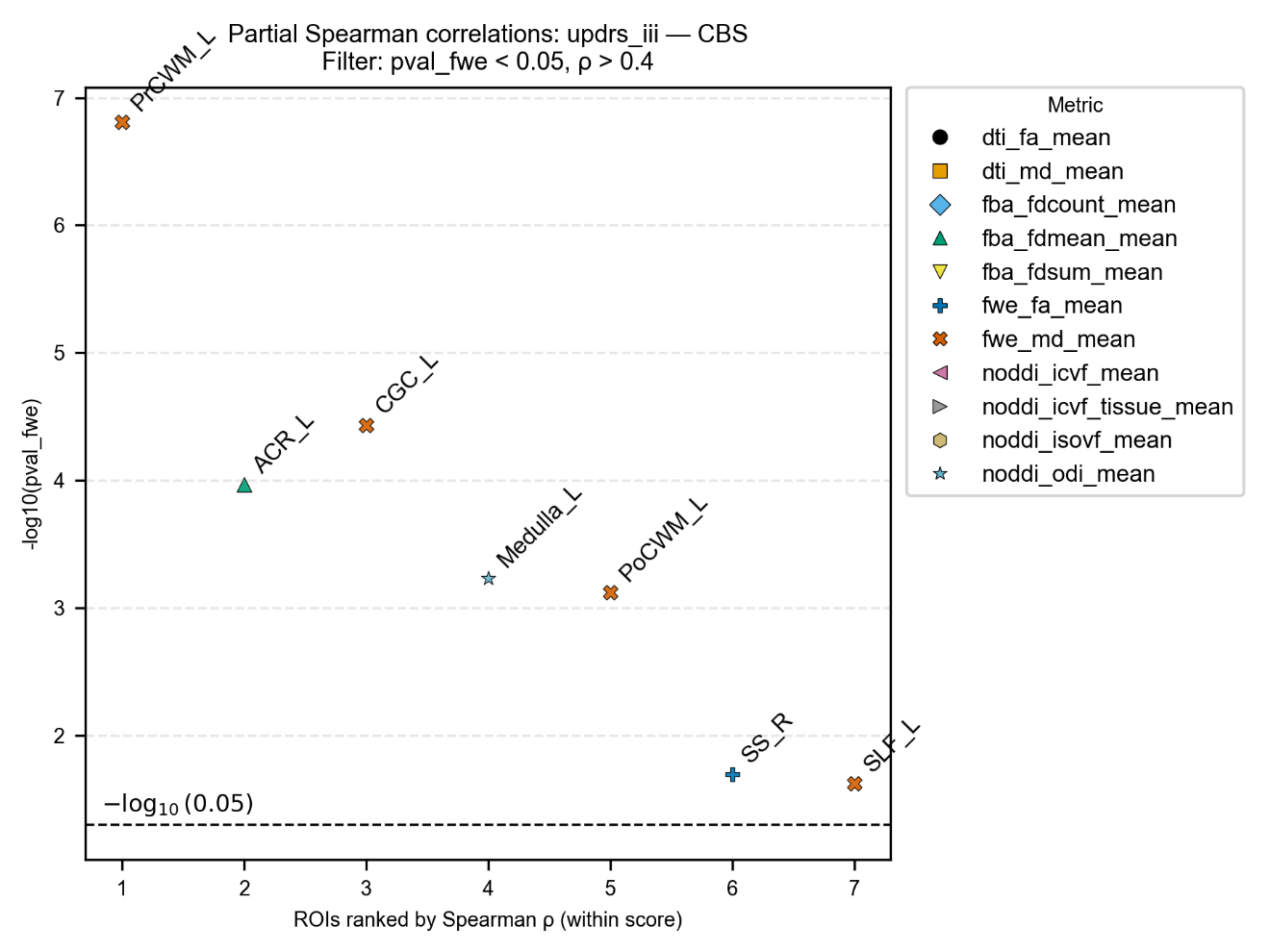

Figure S17 Correlating with MDS-UPDRS III in CBS group.

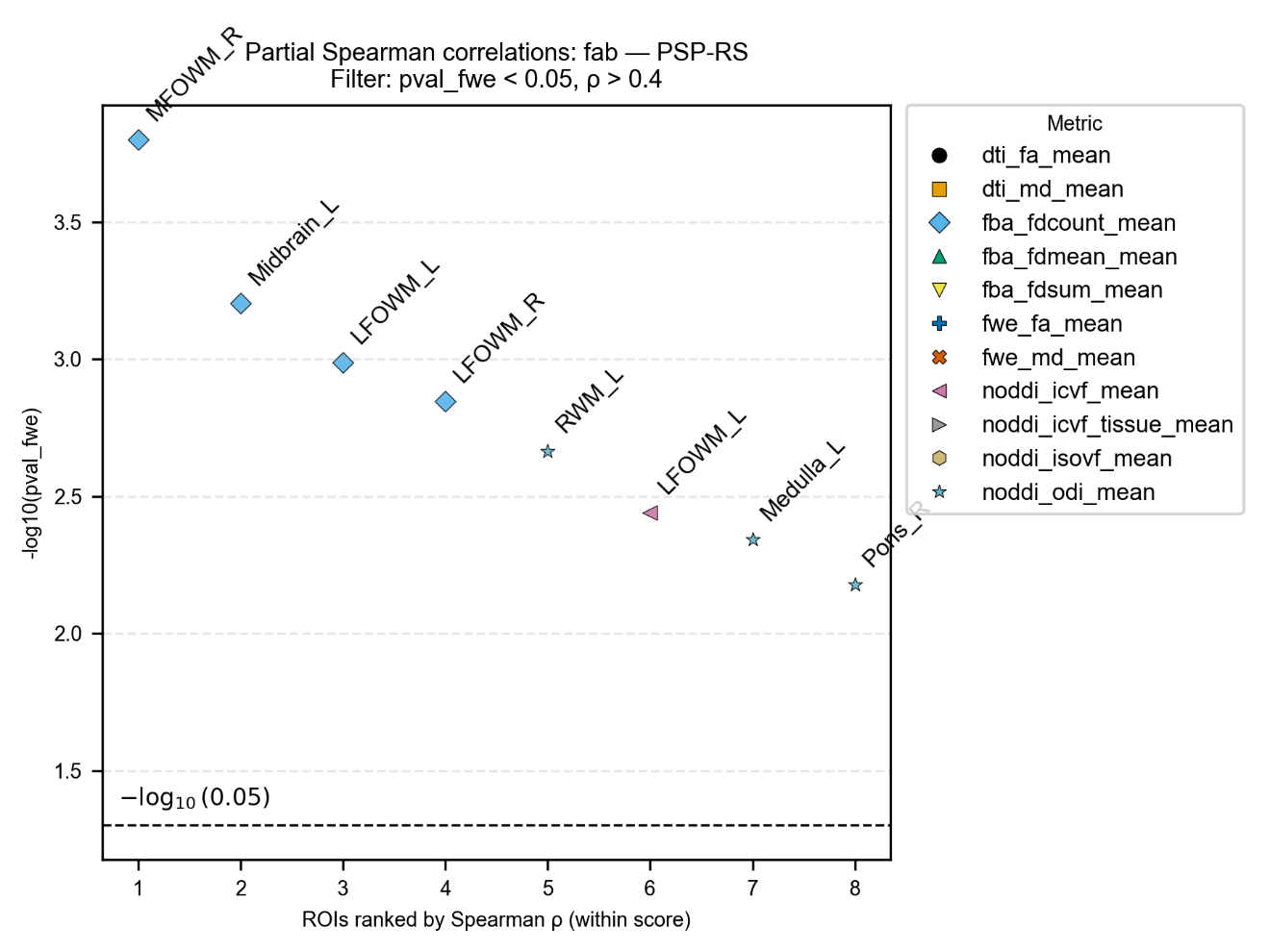

Figure S18 Correlating with FAB in PSP-RS group.

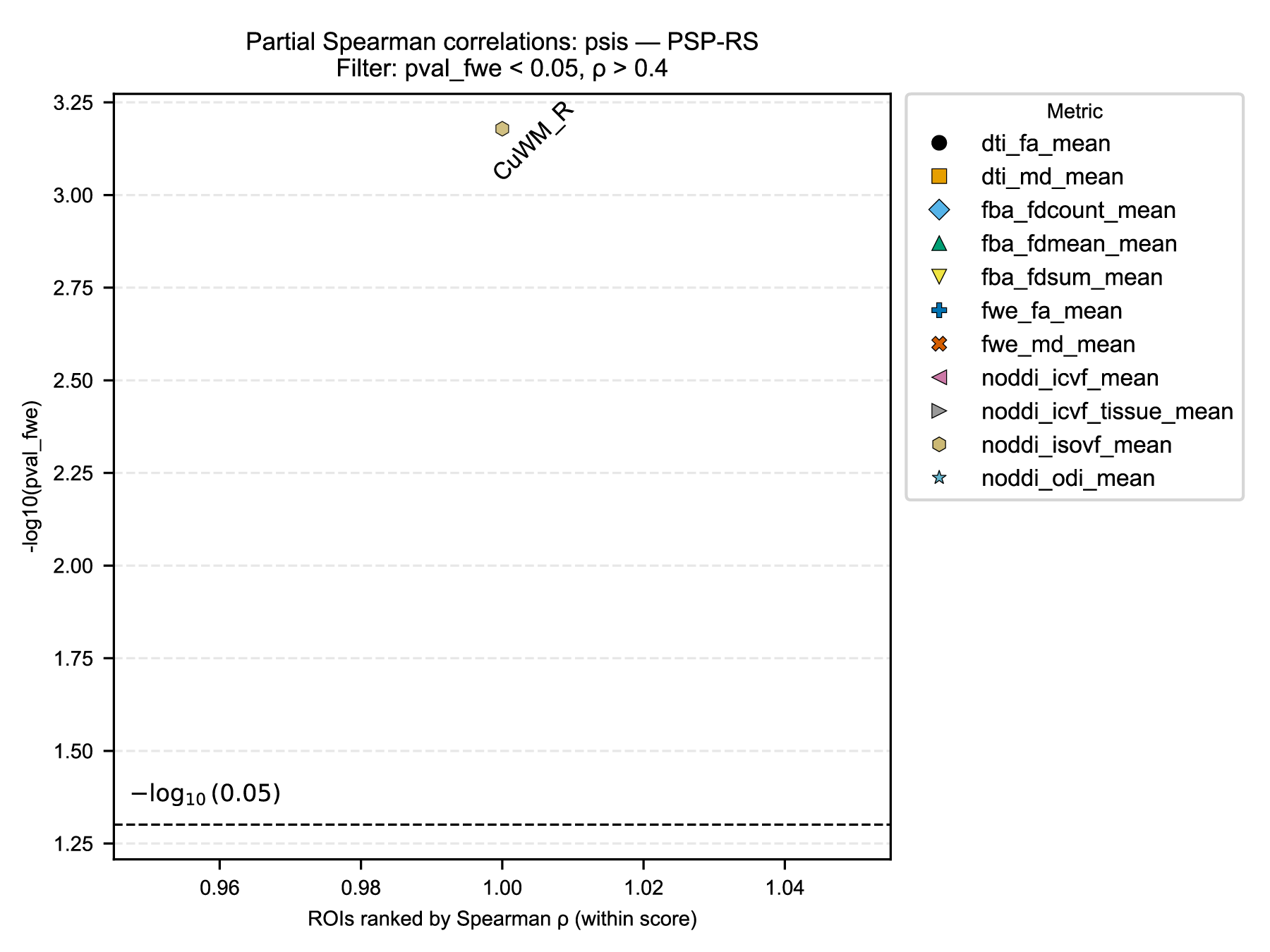

Figure S19 Correlating with PSIS in PSP-RS group.

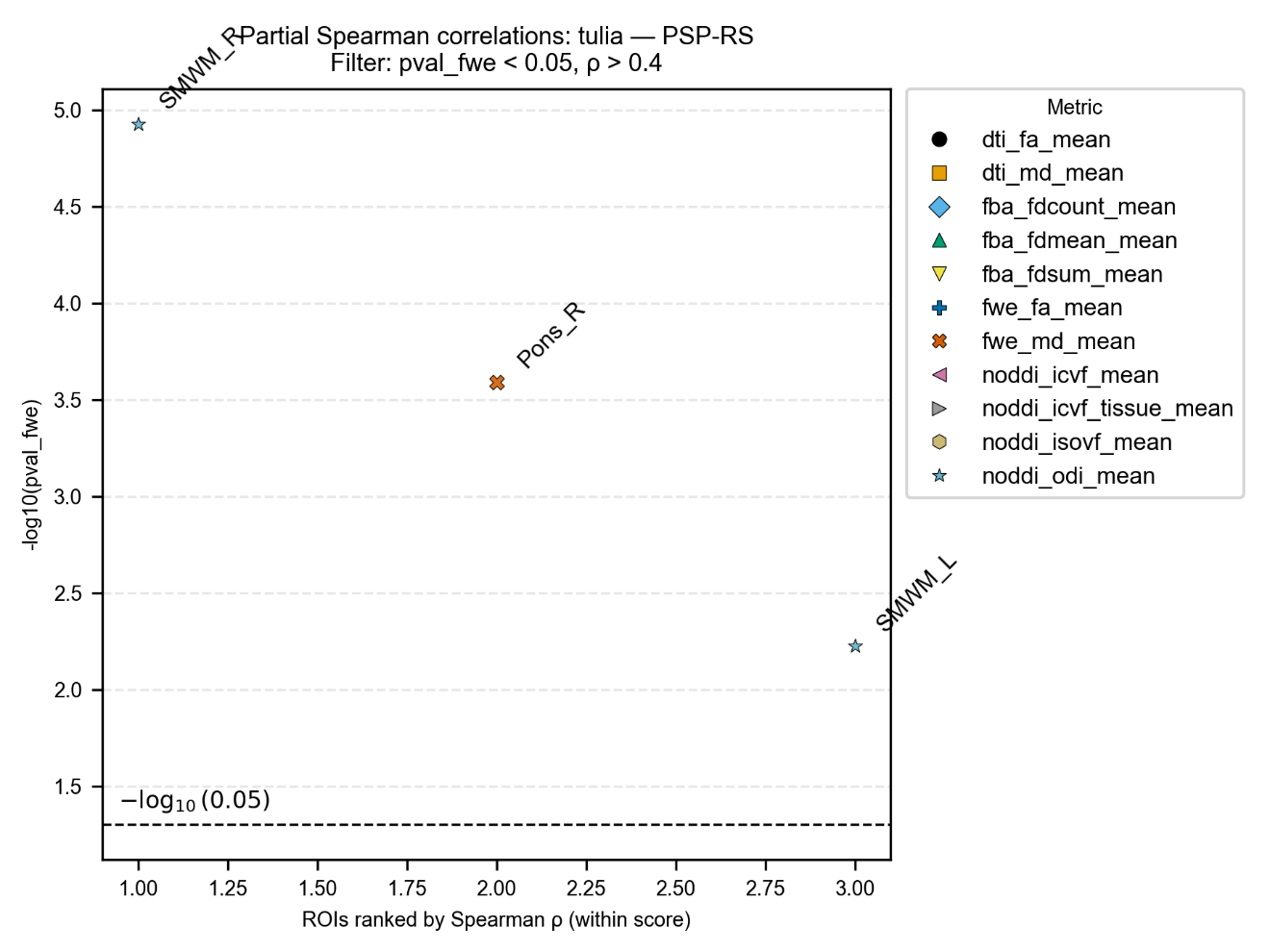

Figure S20 Correlating with TULIA in PSP-RS group.

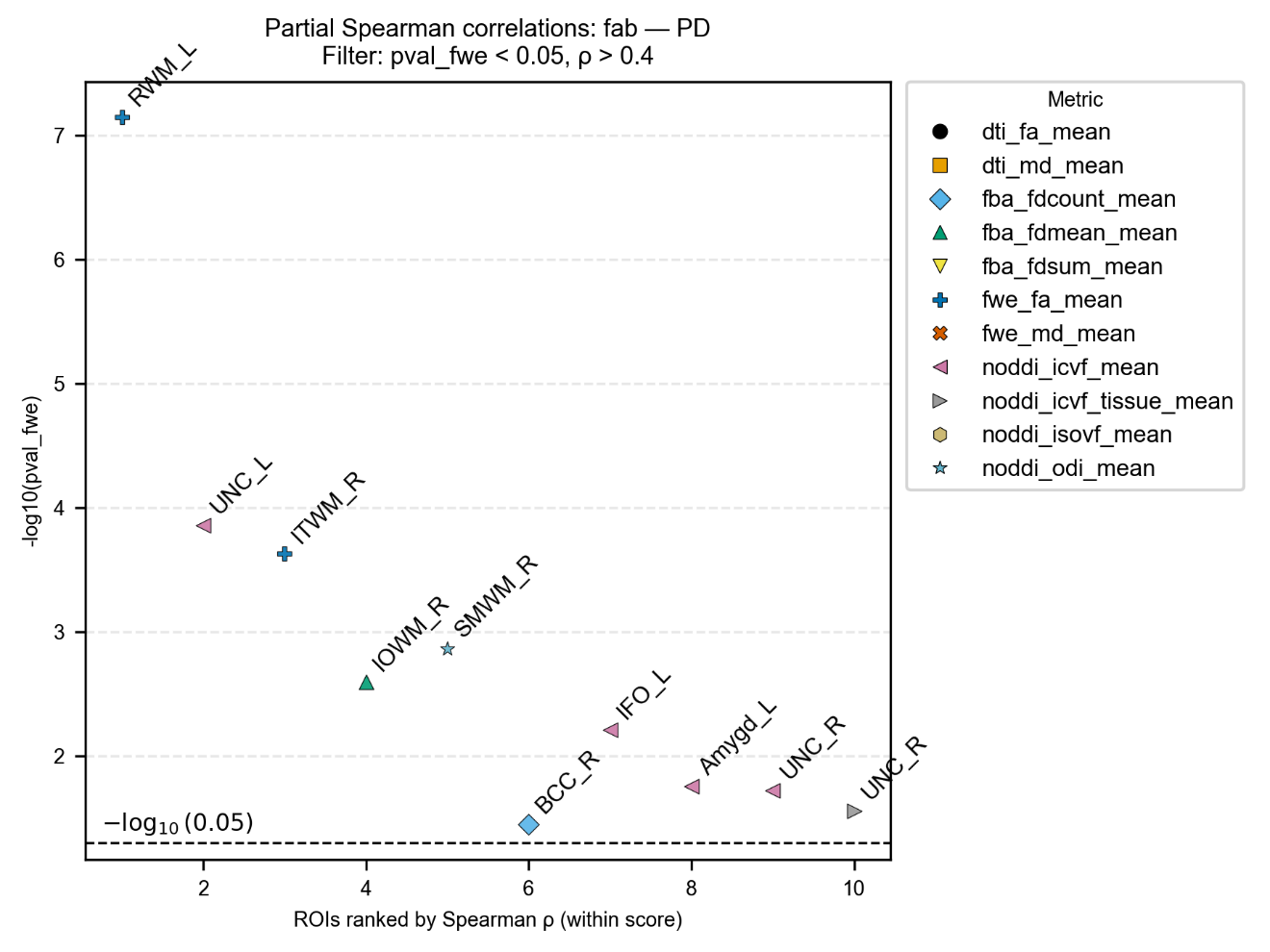

Figure S21 Correlating with FAB in PD group.

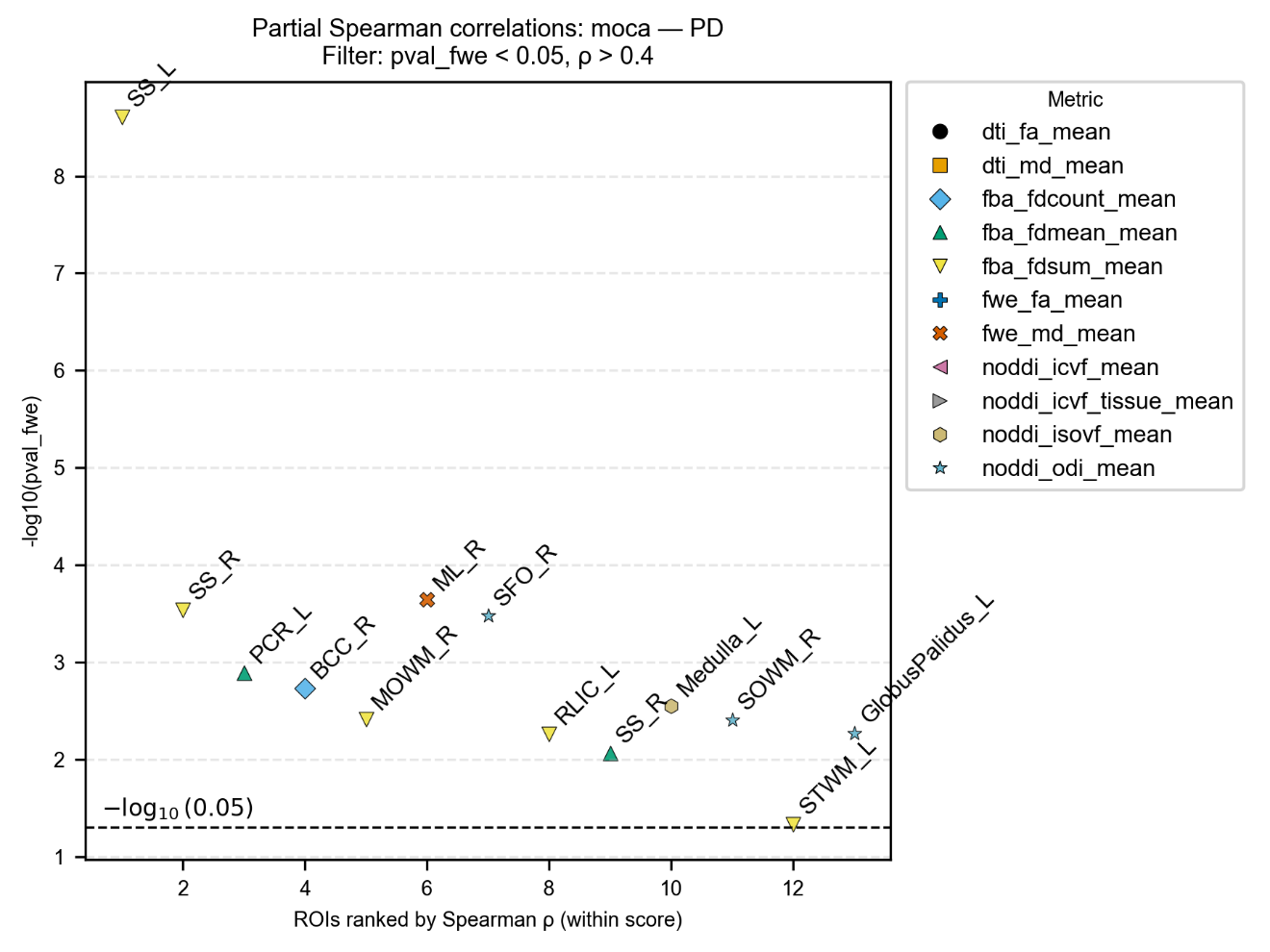

Figure S22 Correlating with MoCA in PD group.
